# Clinical Immunoprofiling Reveals that High Numbers of CD8^+^ and PD-1^+^ Cells Predict Superior Patient Survival Across Major Cancer Types Independent of Major Risk Factors

**DOI:** 10.1101/2024.12.15.24318012

**Authors:** Joao V Alessi, James R Lindsay, Anita Giobbie-Hurder, Bijaya Sharma, Kristen Felt, Priti Kumari, Tali Mazor, Ethan Cerami, William Lotter, Jennifer Altreuter, Jason Weirather, Ian Dryg, Katharina Hoebel, Michael Manos, Elio Adib, Jennifer D. Curtis, Biagio Ricciuti, Alessandro Di Federico, Fatme Ghandour, Eddy Saad, Xin-an Wang, Federica Pecci, Marta Holovatska, Malini M. Gandhi, Melissa E. Hughes, Tess A. O’Meara, Sabrina J. Chan, Kathleen Pfaff, Panagiotis A. Konstantinopoulos, F. Stephan Hodi, Margaret A. Shipp, Sabina Signoretti, Toni Choueiri, Xiao X. Wei, Sandro Santagata, Glenn J. Hanna, Nancy U. Lin, Sara M. Tolaney, Joyce Liu, Peter K. Sorger, Neal Lindeman, Lynette M. Sholl, Jonathan A. Nowak, David Barbie, Mark M. Awad, Bruce E. Johnson, Scott J. Rodig

## Abstract

**Background:** Numerous retrospective studies have shown associations between the number of intratumoral immune cells and patient outcomes for individual cancers treated with specific therapies. However, the clinical value of using a digital pathology platform to enumerate intratumoral immune biomarkers prospectively in the pan-cancer setting has not been established.

**Methods:** We developed ImmunoProfile, a clinical workflow combining automated multiplex immunofluorescence tissue staining, digital slide imaging, and machine learning-assisted scoring to quantify intratumoral CD8^+^, PD-1^+^, CD8^+^PD-1^+^, and FOXP3^+^ immune cells and PD-L1 expression in formalin-fixed, paraffin-embedded tissue samples in a standardized and reproducible manner. Over three years, we prospectively applied ImmunoProfile to biopsies collected from 2,023 unselected cancer patients in the clinical laboratory. We correlated the results with patient survival.

**Results:** In the pan-cancer cohort, patients with intratumoral CD8^+^ or PD-1^+^ cells in the top or middle tertiles had significantly lower risks of death than those in the bottom (CD8^+^: (high vs. low) HR 0.62 [95% CI 0.48 – 0.81], (middle vs. low) HR 0.82 [95% CI 0.67 - 0.99], Wald p=0.002]; PD-1^+^: (high vs. low) HR 0.65 [95% CI 0.51 - 0.83], (middle vs. low) HR 0.74 [95% CI: 0.60 - 0.90], p=0.0009) after controlling for risk factors, including cancer type. In subset analyses, patients with high intratumoral CD8^+^, PD-1^+^, and/or CD8^+^PD-1^+^ cells had lower risks of death from non-small cell lung, colorectal, breast, esophagogastric, head and neck, pancreatic, and ovarian cancers after controlling for clinical risk factors, including stage, and despite distinct therapies (all p ≤ 0.05).

**Conclusions:** Enumerating intratumoral CD8^+^ and PD-1^+^ cells with a clinically validated digital pathology platform predicts patient survival across cancer types independent of clinical stage and despite disparate therapies.

## INTRODUCTION

The foundation for cancer diagnoses remains the visual interpretation of tissue histomorphology using hematoxylin and eosin (H&E)- stained formalin-fixed paraffin-embedded (FFPE) tissue sections. Histomorphologic findings are supplemented with immunohistochemical staining (IHC) results and targeted genetic testing to finalize tumor subclassification, staging, and therapy.^1–4^

The tumor microenvironment (TME) sampled in FFPE tissue sections contains many architectural, cellular, and molecular features that pathologists do not report despite their potential clinical importance.^5^ Of these, markers of anti-tumor immunity are of the greatest interest.^6, 7^ Many retrospective research studies have found that the density of tumor-infiltrating lymphocytes (TILs) and specific immune cell subtypes are associated with outcomes within and across cancer types.^8–10^ These include differential outcomes according to the numbers of intratumoral CD8^+^ and PD-1^+^ T cells, which mark an active immune response, and FOXP3^+^ T cells, which promote immune suppression.^7^ Nevertheless, extending these observations to routine clinical practice has not occurred, primarily due to the substantial time and effort required to enumerate immune cells using traditional, manual methods.^11, 12^

An exception is clinical testing for PD-L1, a PD-1 ligand that enforces T-cell exhaustion.^13^ PD-L1 expression in the TME is associated with a positive patient response to PD-1/PD-L1 inhibitors, and PD-L1 immunohistochemistry (IHC) is routinely performed in the clinical setting.^14^ However, pathologists score PD-L1 IHC subjectively, so its assessment, accuracy, and reproducibility have been questioned.^15, 16^

Automated multiplexed immunostaining, digital imaging, and image analysis platforms have the potential to facilitate, improve, and standardize the reporting of critical immune biomarkers in tissue samples. However, before new tests are used in clinical practice, they must conform to rigorous clinical laboratory standards and undergo clinical validation, ideally with prospective trials.^17–20^ Only Immunoscore, a semi-automated platform that quantifies CD3^+^ and CD8^+^ T cells in tissue sections, has fulfilled these criteria.^21–24^ Immunoscore provides prognostic information not captured by traditional diagnostic and staging criteria for colorectal cancer (CRC) patients and serves as an adjunct to the traditional TNM staging system. However, Immunoscore has not been extended to cancer types other than CRC.

Here, we leveraged advances in automated multiplex immunofluorescence (mIF) staining,^25–27^ digital whole slide image (WSI) capture,^28–30^ and machine learning (ML)-based automated scoring^31, 32^ to develop and validate a new workflow, termed ImmunoProfile, for a clinical laboratory. We used ImmunoProfile to prospectively capture and quantify four key biomarkers associated with promoting (CD8, PD-1) and suppressing (FOXP3, PD-L1) anti-tumor immunity in FFPE tissues acquired from 2,023 patients presenting to a tertiary care cancer center over 3.25 years. We find that we can reliably and quantitatively capture and report multiple key components of the TME in a standardized format. The intratumoral CD8^+^ and PD-1^+^ cell counts predicted patient overall survival (OS) across major cancer types independent of stage and despite diverse therapies.

## METHODS

### Clinical workflow and validation

ImmunoProfile was validated within a CLIA-certified molecular diagnostics laboratory (**Figure 1A**, *Supplementary Figure 1, Supplementary Methods*). FFPE tissue blocks from consented patients (DFCI 11-104; 17-000) were reviewed to select the “best block” for analysis. An H&E-stained slide was used to confirm tumor content. An additional slide was used for automated multiplex immunofluorescence (mIF) staining with a Leica BOND Rx and imaged on an Akoya PhenoImager at 200X magnification.^30, 33, 34^ A minimum of three and a maximum of six regions of interest (ROIs) were selected per whole slide image (WSI) with Inform software (version 2.4.8) according to pre-determined metrics to ensure the capture of immunological “hot spots” (if present). Technicians trained a random-forest-based machine-learning (ML) algorithm in Inform with representative cells and then used it to quantify defined cell types across ROIs. Custom tools (https://github.com/jason-weirather/pythologist) were used to calculate CD8^+^, PD-1^+^, CD8^+^PD-1^+,^ and FOXP3^+^ cell densities, PD-L1 tumor proportion scores (TPS), and PD-L1 combined proportion scores (CPS, *Supplementary Methods*).^35, 36^ An expert pathologist reviewed and signed out each case.

**Figure 1.**
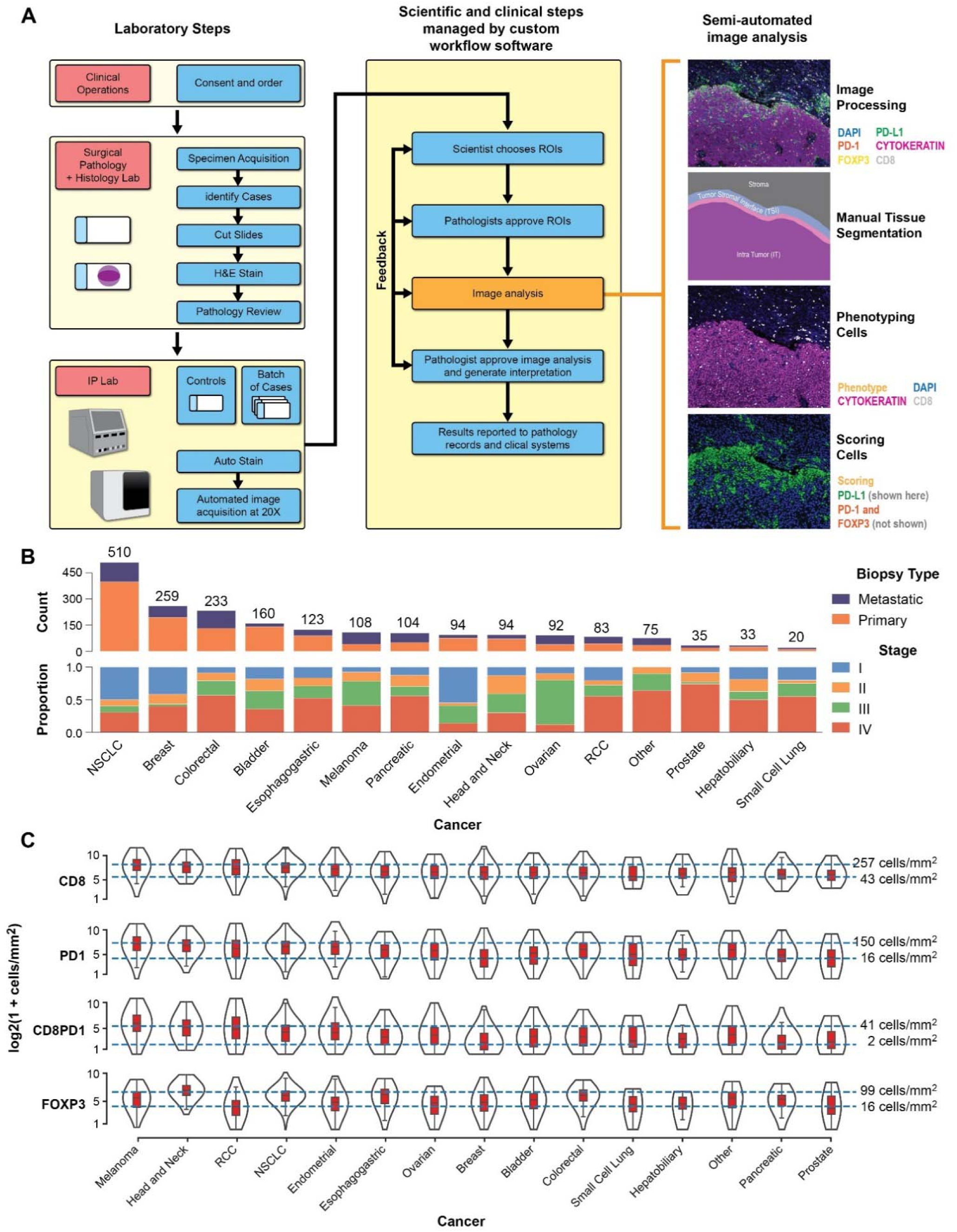
**A:** Schematic outlining the ImmunoProfile workflow in the CLIA-certified laboratory. Consented patients and their diagnostic pathology material were first identified. Slides were cut to generate an H&E-stained slide, which a pathologist reviewed and confirmed to contain diagnostic material. An additional slide was stained on an autostainer and scanned, and a multiplex digital image of the stained tissue was generated. Technicians digitally annotated the images, including selecting regions of interest (ROI) and demarcating the tumor border under pathologists’ review and approval. This was followed by algorithmic training to identify positive and negative staining cells for each biomarker and automated quantitative scoring using image analysis software. A pathologist approved and signed out the image analysis results and the final scores. Results were reported to a clinically accessible database. Images and image analysis results were retained in a HIPAA-compliant data warehouse. Further details are provided in *Supplementary Methods*. **B:** The 2,023-patient pan-cancer ImmunoProfile cohort. The cases are separated into 14 major diagnostic categories and “other” rare cancer types. The number of cases within each major cancer type is further annotated by the tissue acquisition site (primary versus metastatic) and patients’ clinical stage. Further details are provided in *Supplementary Materials*. **C:** Violin plot showing the distributions of intratumoral immune cell density (log_2_ of cells per mm^2^) for all cases, separated according to cancer type and organized from the highest to lowest average CD8^+^ immune cells. The blue lines indicate the immune cell densities at the 75^th^ and 25^th^ percentiles for the pan-cancer cohort. The black lines represent the mean value, the red boxes represent the 75^th^ and 25^th^ percentile thresholds, and the black outlines represent case numbers at the indicated density for each cancer type. Note that there are cases within the highest and lowest quartiles for each biomarker for each cancer type.

### Clinical Annotation and Survival Analysis

Patient information was obtained from medical records and the DFCI OncDRS data warehouse with IRB approval.^37^ Appropriate clinical experts also manually reviewed every case (*Supplementary Table 1*).

### Statistical Methods and Prognostic Models

Comparisons between groups for continuous scaled data used Wilcoxon rank-sum or Kruskal-Wallis tests. Categorical comparisons used Fisher’s exact or chi-squared tests. Statistical significance was p ≤ 0.05. Overall survival (OS) was determined from the date of tissue acquisition and death from any cause. The follow-up of patients alive at data retrieval was censored at the date of last contact. We divided patients by their respective biomarker scores into tertiles when there were >100 cases and ≥50 deaths for a cancer type. We split patients by their respective biomarker scores at the median for smaller cohorts.

OS was estimated using the Kaplan-Meier method and compared using log-rank testing; 95% confidence intervals were calculated using log(-log) methodology. Median follow-up was based on Kaplan-Meier estimation with an inverted censor.

Associations between biomarker measurements and OS were assessed using Cox proportional hazards models adjusted for clinical risk factors selected before analyses, including cancer type (for the pan-cancer analysis) and the American Joint Committee on Cancer (AJCC) stage.^38^, age, alcohol use, smoking, and sex. To avoid overfitting, the number of risk factors included in each model was adjusted according to cohort size and deaths before analysis. Multivariable models were fit for cancer types with ≥ 30 deaths only. Immune cells with adjusted Wald p-values ≤ 0.1 from the single marker analyses were considered candidates in the multivariable setting (see *Supplementary Methods*).

## RESULTS

### ImmunoProfile reliably quantifies key immunological biomarkers in clinical samples

We modified a research protocol^35^ to develop a new clinical workflow, ImmunoProfile, which combines automated mIF tissue staining, digital WSI capture, and ML-directed analysis to calculate CD8^+^, PD-1^+^, CD8^+^PD-1^+^, FOXP3^+^ cell densities and PD-L1 TPS and CPS in FFPE tissue sections in a CLIA-certified laboratory (**Figure 1A**). Validation studies demonstrated a high concordance between ImmunoProfile and IHC (*Supplementary Figure 1*). The application of ImmunoProfile to clinical samples revealed minimal run-to-run variability in staining, imaging, and scoring over time (*Supplementary Figure 2*). We observed minimal variation in results across technicians and pathologists, and < 3% of cases failed for technical reasons (*Supplementary Methods*). The entire multistep workflow met or exceeded clinical laboratory quality-control requirements and was approved for prospective clinical application.

### Patients with high intratumoral immune cells and PD-L1 expression are found across cancer types

In a prospective study, we analyzed biopsy and resection tissue specimens sent for pathology review from all consented cancer patients for 3 years. The final 2,023 case-cohort included 14 major cancer types, biopsies from primary and metastatic sites, and patients with low- and high-stage disease, as expected for a tertiary cancer center (**Figure 1B**, *Supplementary Table 1*).

We found that intratumoral immune cell numbers and PD-L1 expression varied substantially across cases and cancer types (**Figure 1C**, *Supplementary Figure 3*). When we divided cases by their respective biomarker scores into the cohort’s highest, middle, and lowest tertiles, we found cases with individual scores in the top and bottom tertiles for every cancer type (*Supplementary Figure 3*). Thus, patients can have “hot” or “cold” tumors irrespective of cancer type, although patients with some cancer types are more likely to have a “hot” tumor microenvironment than others.

### Immune biomarker scores split by tertiles stratify patient survival in pan-cancer analyses

We next examined the prognostic significance of the reported immune biomarker scores. In a pan-cancer analysis (n = 2,023, 632 deaths), we divided patients by their respective numbers of intratumoral CD8^+^, PD-1^+^, CD8^+^PD-1^+^, and FOXP3^+^ cells into tertiles (*Supplementary Table 2*). Those in the top tertiles had longer OS than those in the middle, which, in turn, had longer OS than those in the bottom tertile for each immune cell type (all log-rank p ≤ 0.0001, **Figure 2A**). Patients whose tumors had PD-L1 combined proportion scores (CPS) in the highest and middle tertiles had longer OS than those patients with tumors in the lowest tertile (p = 0.002, *Supplementary Figure 4A*). In contrast, patients whose tumors had PD-L1 tumor proportion scores (TPS) in the highest, middle, or lowest tertiles had similar OS (*Supplementary Figure 4B*). These data reveal that intratumoral CD8^+^, PD-1^+^, CD8^+^PD-1^+^, and FOXP3^+^ cell densities and PD-L1 CPS captured in FFPE tissues by this clinical test predict survival in the pan-cancer setting.

**Figure 2.**
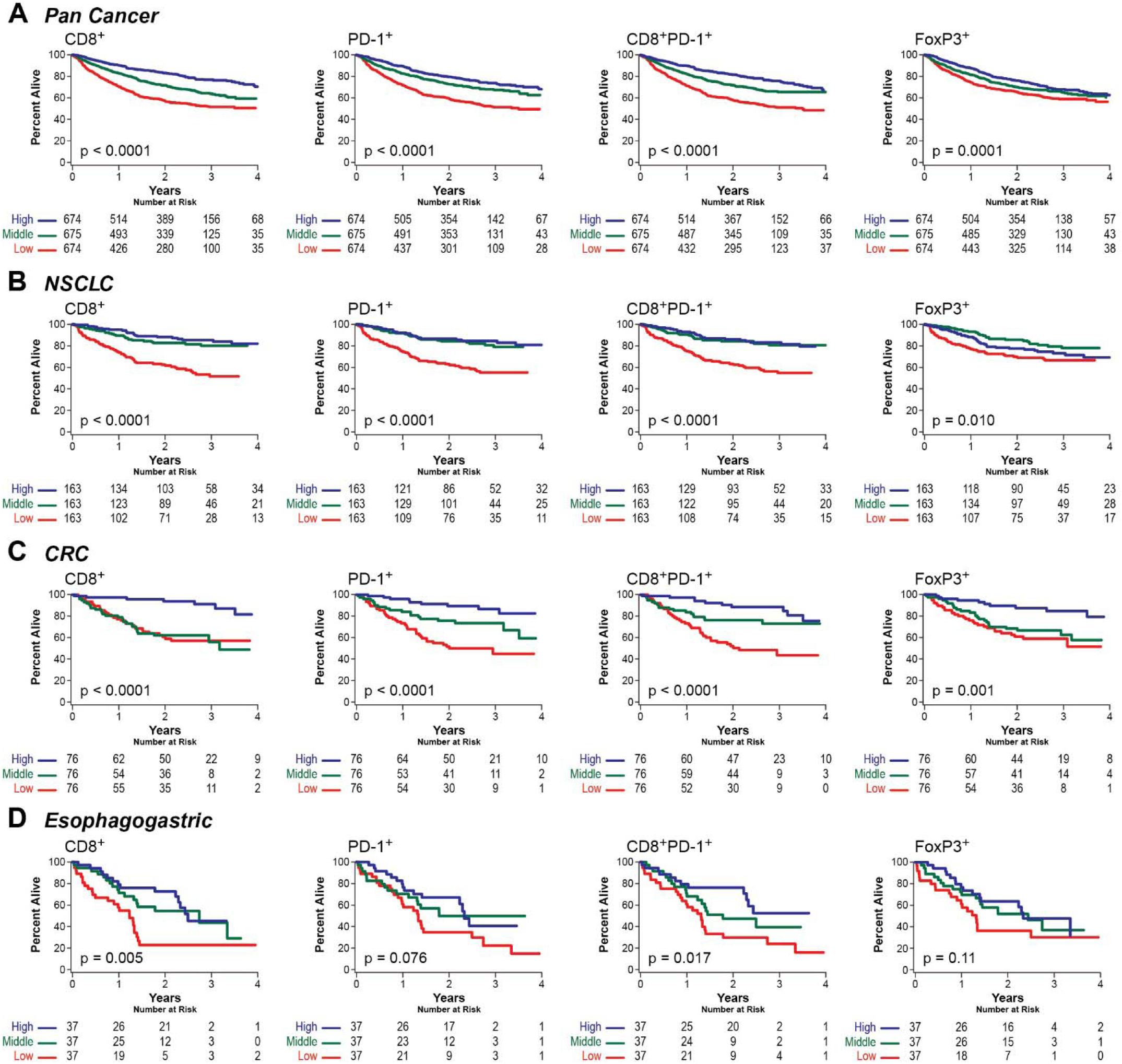
Kaplan-Meier estimates of overall survival (OS) with biomarker values divided into tertiles. The pan-cancer and major cancer sub-types were divided according to their CD8^+^, PD-1^+^, CD8^+^PD-1^+^, and FoxP3^+^ immune cell densities into high (blue), middle (green), and low (red) tertiles for each respective group. The number of patients at risk of death at the indicated time from diagnosis is indicated for **A:** all patients (n = 2,023), and patients with **B:** non-small cell lung cancer (NSCLC, n = 489), **C:** colorectal cancer (CRC, n = 228), and **D:** esophagogastric cancer (EGD, n = 111), The p-values by the log-rank test are indicated.

### Immune biomarker scores split into tertiles predict OS for select major cancer types

We next divided patients with non-small cell lung cancer (NSCLC, n = 489), colorectal cancer (CRC, n = 228), esophagogastric cancer (EGC, n = 111), pancreatic cancer (PAN, n = 104), and bladder cancer (BLAD, n= 159) into high, middle, and low groups based on tertile divisions of their respective biomarker scores (*Supplementary Table 2*). Those with NSCLC, CRC, and EGC and intratumoral CD8^+^, PD-1^+^, and/ or CD8^+^PD-1^+^ cells in the highest tertiles for each group had significantly better OS than those in the middle or lowest tertiles (all significant log-ranks p ≤ 0.05, **Figure 2B-D**). Patients with PANC and PD-1^+^ cells in the highest and lowest tertiles had better OS than those with PD-1^+^ cells in the middle tertile (p =0.03, *Supplementary Figure 5A*), which may reflect an unusual immune response profile or a weaker association between intratumoral immune cells and OS for this cancer type.^39^ Patients with BLAD did not show differences in OS when cases were split in this manner (all p > 0.05, *Supplementary Figure 5B*). None of the groups showed significant differences in OS when split by PD-L1 CPS and TPS scores (all p > 0.05).

Given the large number of patients with NSCLC, we further split patients into those receiving immune checkpoint blockade (ICB, anti-PD-1, or anti-PD-L1, n = 114) or not (no-ICB, n = 372). For both groups, we found that those with CD8^+^, PD-1^+^, or CD8^+^PD-1^+^ cells in the highest tertiles had longer OS than those in the lowest tertiles (all p < 0.001, **Figure 3A-B**). Thus, intratumoral CD8^+^, PD-1^+^, CD8^+^PD-1^+^ and/or FOXP3^+^ cell numbers are associated with OS among patients with NSCLC, CRC, EGC, and PANC by this analysis.

**Figure 3.**
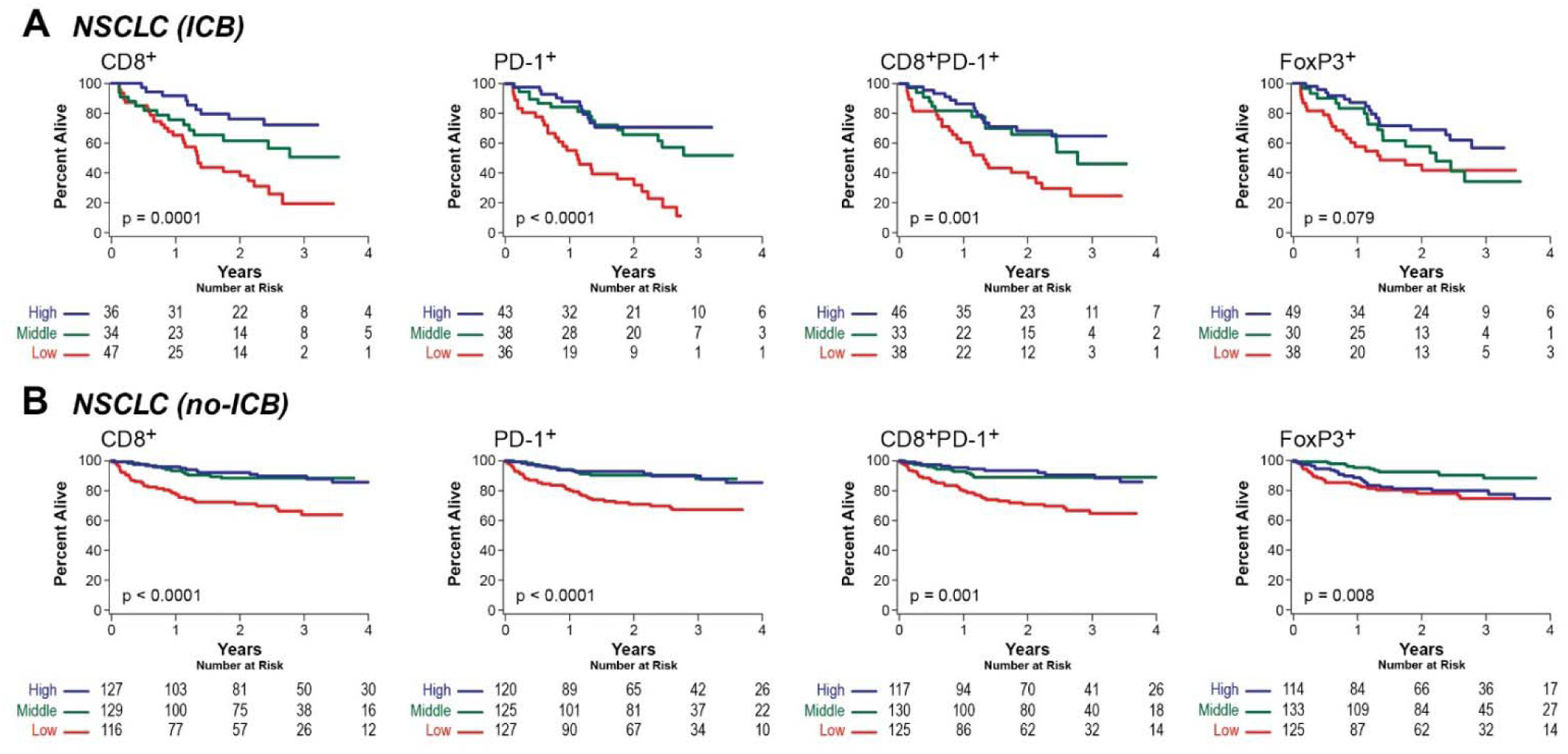
Kaplan-Meier curves of overall survival (OS) according to tertiles of the biomarker distributions. The patients were divided according to their CD8^+^, PD-1^+^, CD8^+^PD-1^+^, and FoxP3^+^ immune cell densities into high (blue), middle (green), and low (red) tertiles for each respective group. The number of patients at risk at the indicated number of years from the time of diagnosis is indicated for **A:** Patients with NSCLC treated with immune checkpoint blockade (NSCLC (ICB), n = 117), **B:** Patients with NSCLC treated without immune checkpoint blockade (NSLCL (no-ICB), n = 372). The p-values by the log-rank test are indicated.

### Immune biomarker scores split at the median predict OS for select major cancer types

We had fewer than 100 patients and/or 50 deaths for patients with hormone-receptor-positive/ HER2/neu-negative breast cancer (HR^+^/HER2^-^ BRCA, n = 168), estrogen/progesterone/ HER2/neu negative breast cancer (TN BRCA, n = 55), ovarian cancer (OVCA, n = 90), head and neck squamous cell cancer (HNSCC, n = 88), renal cell carcinoma (RCC, n = 79), endometrial cancer (ENDOM, n = 89), and cutaneous melanoma (MEL, n = 98). Therefore, we divided these patients into “high” and “low” groups based on the median values of their respective biomarker scores. Patients with HR^+^/HER2^-^ BRCA, TN BRCA, and OVCA and high intratumoral CD8^+^ or CD8^+^PD-1^+^ cell numbers had significantly longer OS than those with low numbers (each significant log-rank p < 0.05, **Figure 4A-C)**. Patients with HNSCC and high CD8^+^PD-1^+^ or FOXP3^+^ cell numbers tended toward more favorable OS (p = 0.08 and 0.06, **Figure 4D**). Patients with RCC, ENDOM, or cutaneous MEL did not show significant differences in OS by this analysis (all p > 0.05).

**Figure 4.**
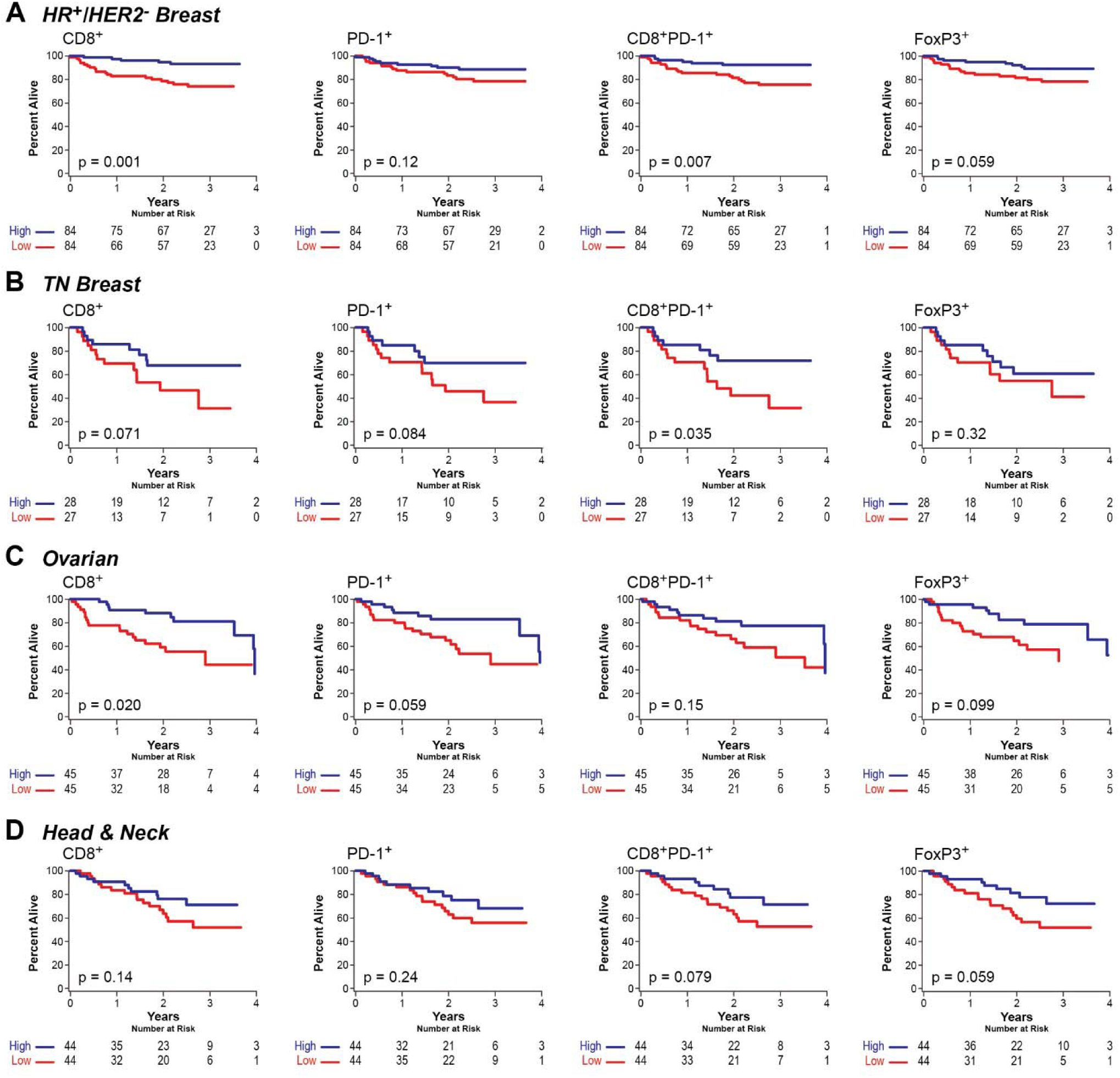
Kaplan-Meier estimates of overall survival (OS) for biomarker values divided at the median. The major cancer sub-types were divided according to their CD8^+^, PD-1^+^, CD8^+^PD-1^+^, and FoxP3^+^ immune cell densities into high (blue) and low (red) groups relative to each cancer type’s median values. The number of patients at risk of death at the indicated time from diagnosis is indicated for patients with **A:** hormone receptor-positive/HER2 negative breast cancer (HR+/HER2^-^ Breast, n = 168), **B:** triple-negative breast cancer (TN Breast, n = 55), **C:** ovarian carcinoma (n = 90), and **D:** head & neck squamous cell carcinoma (n = 88). The p-values by the log-rank test are indicated.

We next divided patients according to their PD-L1 scores. Those with HR^+^/HER2^-^ BRCA and high CPS had longer OS than those with low CPS (p = 0.002; *Supplementary Figure 6A*). In contrast, patients with RCC and *high* TPS had significantly *shorter* OS than those with low TPS (p = 0.049; *Supplementary Figure 6B*). Given the low number of patients (35 or less) with additional cancer types (prostate, hepatobiliary, small cell lung cancer), we did not perform survival analyses for these groups. Thus, CD8^+^ and/or CD8^+^PD-1^+^ cell numbers predict the survival of patients with HR^+^/HER2^-^ BRCA, TN BRCA, and OVCA. In contrast, PD-L1 TPS predicts the survival of patients with RCC.

### Multiple biomarkers predict OS after adjustments for stage and other risk factors

We next examined whether individual biomarkers predicted OS after accounting for major clinical risk factors, including cancer type, biopsy site, AJCC stage, age, gender, smoking, and alcohol use (**Table 1***; Supplementary Figure 7*). Importantly, we selected the risk factors used in each cancer-specific model before calculating the results to avoid overfitting (See *Methods*).

**Table 1.** Biomarker Associations with Overall Survival from Univariate Cox Models with Clinical Risk Factor Adjustments.

| Cancer Type | Patients/ deaths | Division | p-value |  |  |  | Risk Factor Adjustments |
| --- | --- | --- | --- | --- | --- | --- | --- |
|  |  |  | CD8<sup>+</sup> | PD-1<sup>+</sup> | CD8+PD-1<sup>+</sup> | FOXP3<sup>+</sup> |  |
| Pan-cancer | 2023 / 632 | Tertile | **0.0001** | **0.0001** | **0.0001** | **0.0001** | Cancer type, primary/met, smoking, alcohol, age, gender |
| NSCLC (all) | 489 / 115 | Tertile | **0.009** | **0.001** | **0.0001** | 0.31 | AJCC stage, smoking, alcohol, age, gender |
| NSCLC (ICB) | 117 / 54 | Tertile | **0.001** | **0.0002** | **0.04** | **0.03** | AJCC stage, smoking, alcohol, age, gender |
| NSCLC (No-ICB) | 372 / 61 | Tertile | 0.07 | 0.08 | **0.003** | 0.19 | AJCC stage, smoking, alcohol, age, gender |
| CRC | 228 / 66 | Tertile | **0.006** | **0.0006** | **0.006** | 0.27 | AJCC stage, MSI status, smoking, alcohol, age |
| PANC | 104 / 71 | Tertile | 0.23 | **0.007** | 0.48 | **0.04** | AJCC stage, smoking, alcohol, age |
| EGC | 111 / 56 | Tertile | **0.02** | 0.12 | 0.08 | 0.36 | AJCC stage, smoking, alcohol, age |
| BLAD | 159 / 57 | Tertile | 0.69 | 0.80 | 0.75 | **0.03** | AJCC stage, smoking, alcohol, age, gender |
| HR+/HER2 BRCA | 168 / 25 | Median | **0.002** | 0.24 | **0.02** | **0.05** | Low (I/II) or high (III/IV) stage |
| TN BRCA | 55 / 21 | Median | 0.35 | 0.24 | 0.06 | 0.78 | Low (I/II) or high (III/IV) stage |
| HNSCC | 88 / 27 | Median | 0.14 | 0.27 | **0.05** | 0.26 | Low (I/II) or high (III/IV) stage |
| OVCA | 90 / 30 | Median | **0.02** | 0.06 | 0.09 | **0.05** | Primary vs metastatic disease |
| RCC | 79 / 24 | Median | 0.97 | 0.96 | 0.25 | 0.90 | AJCC stage |
| ENDOM | 89 / 19 | Median | 0.35 | 0.30 | 0.18 | 0.43 | Primary vs metastatic disease |
| Cut. MEL | 98 / 18 | Median | 0.52 | 0.45 | 0.54 | 0.83 | AJCC stage |

NSCLC= non-small cell lung cancer, ICB= treatment with immune checkpoint blockade, no-ICB= no treatment with checkpoint blockade, CRC= colorectal carcinoma, PANC= pancreatic carcinoma, EGC= esophagogastric carcinoma, BLAD= bladder carcinoma, HR<sup>+</sup>/ HER2<sup>-</sup> BRCA= hormone-receptor-positive, Her2-negative breast carcinoma, TN BRCA= triple-negative breast carcinoma, HNSCC= head and neck squamous cell carcinoma, OVCA= ovarian carcinoma, RCC= renal cell carcinoma, ENDOM= Endometrial carcinoma; cut. MEL= cutaneous melanoma. AJCC= American Joint Committee on Cancer. MSI= microsatellite instability.

In the pan-cancer analysis, patients with CD8^+^, PD-1^+^, CD8^+^PD-1^+^, or FOXP3^+^ cells in the top tertiles had longer OS than those in lower tertiles after risk factor adjustments (all p = 0.0001, **Table 1**). In sub-group analyses, those with NSCLC, with or without ICB, CRC, EGC, BLAD, HR^+^/HER2^+^ BRCA, HNSCC, and OVCA and high numbers of CD8^+^, PD-1^+^, CD8^+^PD-1^+^, and/or FOXP3^+^ cells had significantly longer OS than those with low numbers after risk factor adjustments (all significant p ≤ 0.05, **Table 1**). Intriguingly, for patients with PANC, the association between intratumoral PD-1^+^ cell numbers and OS was highly significant after risk factor adjustments, including stage (p = 0.007). Patients with TN BRCA and high numbers of CD8^+^PD-1^+^ cells trended towards more prolonged OS after adjusting for stage (p = 0.06, **Table 1**). Patients with RCC, ENDOM, and cutaneous MEL did not show significant differences in OS by this analysis (**Table 1**).

In the pan-cancer analysis, patients divided into tertiles by PD-L1 CPS or TPS did not show significant differences in OS after risk adjustments (*Supplementary Table 3*). However, patients with NSCLC, treated with an immune checkpoint inhibitor (anti-PD-1 or anti-PD-L1), and high PD-L1 CPS (top tertile) or TPS (>1%) had longer OS than those with low PD-L1 scores (p = 0.04, 0.05; *Supplementary Table 3*). Similarly, patients with CRC and high TPS (> 5%) had longer OS than those with low TPS (p = 0.02, *Supplementary Table 3*). In contrast, patients with RCC and TPS above the median for the group had worse OS than those below after adjusting for stage (p = 0.04, *Supplementary Table 3*). We conclude that intratumoral CD8^+^, PD-1^+^, and/or CD8^+^PD-1^+^ cell numbers provide prognostic information not captured by major clinical risk factors for patients with NSCLC, CRC, PANC, EGC, BLAD, HR^+^/HER2^+^ BRCA, HNSCC, and OVCA. PD-L1 scores provide prognostic information not captured by major clinical risk factors for patients with NSCLC treated with ICB and patients with CRC and RCC.

### Multivariable biomarker analyses reveal specific immune cell types most closely associated with OS

Numbers of CD8^+^, PD-1^+^, and CD8^+^PD-1^+^ cells were highly correlated across cases (*r*= 0.8-0.92, *Supplementary Figure 8*). The numbers of FOXP3^+^ cells and PD-L1 scores were less so (*r*= 0.26-0.63, *Supplementary Figure 8*). Therefore, we assessed which individual immune biomarker(s) were most strongly and independently associated with the risk of death using multivariable biomarker modeling (see *Methods*). In the pan-cancer cohort, patients with high numbers of PD-1^+^ or CD8^+^ cells were independently associated with a reduced risk of death after including clinical risk factor adjustments (**Table 2**). Those with PD-1^+^ cells in the highest tertile had a 35% reduced risk of death, and those in the middle tertile had a 26% reduced risk compared to those in the lowest tertile (p= 0.0009). We found similar relationships for CD8^+^ cells (p = 0.002, **Table 2**).

**Table 2.** Risk of Death in Multivariable Biomarker Cox Models with Clinical Risk Factor Adjustments for Cancers with High Case Numbers.

| Cancer Type | Marker | Division | Hazard Ratio of Survival (95% CI) |  | P-value |
| --- | --- | --- | --- | --- | --- |
| Pan Cancer | PD-1 | High vs. Low | 0.65 (0.51 - 0.83) |  | 0.0009 |
|  |  | Middle vs. Low | 0.74 (0.60 - 0.90) |  |  |
| Pan Cancer | CD8 | High vs. Low | 0.62 (0.48 - 0.81) |  | 0.002 |
|  |  | Middle vs. Low | 0.82 (0.67 - 0.99) |  |  |
| NSCLC (all) | CD8 PD-1 | High vs. Low | 0.44 (0.27 - 0.71) |  | 0.0001 |
| NSCLC (all) | CD8 PD-1 | Middle vs. Low | 0.44 (0.28 - 0.70) |  |  |
| NSCLC (ICB) | PD-1 | High vs. Low | 0.22 (0.10 - 0.50) |  | 0.0002 |
| NSCLC (ICB) | PD-1 | Middle vs. Low | 0.33 (0.17 - 0.66) |  |  |
| NSCLC (no ICB) | CD8 PD-1 | High vs. Low | 0.51 (0.24 - 1.05) |  | 0.003 |
| NSCLC (no ICB) | CD8 PD-1 | Middle vs. Low | 0.33 (0.17 - 0.64) |  |  |
| CRC | PD-1 | High vs. Low | 0.20 (0.09 - 0.50) |  | 0.0006 |
| CRC | PD-1 | Middle vs. Low | 0.65 (0.34 - 1.17) |  |  |
| PANC | PD-1 | High vs. Middle | 0.36 (0.19 - 0.68) |  | 0.007 |
| PANC | PD-1 | Low vs. Middle | 0.60 (0.31 - 1.14) |  |  |
| EGC | CD8 | High vs. Low | 0.41 (0.20 - 0.82) |  | 0.02 |
| EGC | CD8 | Middle vs. Low | 0.84 (0.40 - 1.76) |  |  |
| BLAD | FOXP3 | High vs. Middle | 0.40 (0.20 - 0.80) |  | 0.03 |
| BLAD | FOXP3 | Low vs. Middle | 0.70 (0.33 - 1.48) |  |  |
| OVCA | CD8 | High vs. Low | 0.38 (0.18 - 0.84) |  | 0.02 |

NSCLC= non-small cell lung cancer, ICB= treatment with checkpoint blockade, no-ICB= no treatment with immune checkpoint blockade, CRC= colorectal carcinoma, PANC= Pancreatic carcinoma, EGC= esophagogastric carcinoma, BLAD= bladder carcinoma, cut. MEL= cutaneous melanoma; OVCA= ovarian carcinoma. OS= overall survival. The clinical risk factors considered in the multivariable modeling for each group are listed in Table 1.

For patients with NSCLC, intratumoral CD8^+^PD-1^+^ cell numbers best predicted the risk of death after accounting for risk factors, including stage and smoking. Those with CD8^+^PD-1^+^ cells in the highest and the middle tertiles each had a 56% reduced risk of death compared to those in the bottom tertile (p = 0.0001, **Table 2**). For those treated with ICB, intratumoral PD-1^+^ cells best predicted the risk of death; for those not treated with ICB, CD8^+^PD-1^+^ cells best predicted the risk of death (p = 0.003, **Table 2**).

After risk adjustments, including for MSI status, patients with CRC and intratumoral PD-1+ cells in the highest tertile had an 80% reduced risk of death compared with those in the lowest tertile (p= 0.0006, **Table 2**). Patients with EGC and CD8^+^ cells in the highest tertile had a 59% reduced risk compared to those in the lowest (p= 0.02). Patients with OVCA and numbers of CD8^+^ cells above the median had a 62% reduced risk compared with those below (p = 0.02, **Table 2**).

Patients with PANC and BLAD had the most significantly reduced risk of death between those in the highest and middle tertiles for PD-1^+^ and FOXP3^+^ cells, respectively. Those with PANC and intratumoral PD-1^+^ cells in the highest tertile had a 64% reduced risk compared to those in the middle (p = 0.007). Patients with BLAD and FOXP3^+^ cells in the highest tertile had a 68% reduced risk compared to those in the middle (p = 0.03, **Table 2**). Thus, we find that specific immune biomarkers, most commonly PD-1^+^ and CD8^+^ immune cell numbers, best predict patients’ risk of death independent of currently recognized risk factors, including stage, for major cancer types, including NSCLC, CRC, EGC, OVCA, PANC, and BLAD.

### Univariable analyses reveal that specific immune cell populations are most closely associated with OS

We did not attempt multivariable analysis for patients with cancer types having less than 30 deaths. Instead, we described the risk of death with the biomarker most significantly associated with OS after adjusting for clinical risk factors (as shown in **Table 1**). Patients with HR^+^ /HER2^-^ BRCA and high CD8^+^ cell numbers had an 80% lower risk of death than those with low numbers (p = 0.002, **Table 3**). Patients with TN BRCA and high CD8^+^PD-1^+^ cell numbers had a lower risk of death than those with low numbers that approached significance (p = 0.06; **Table 3**). Patients with HNSCC and high CD8^+^PD-1^+^ cell numbers had a 56% lower risk than those with low numbers (p = 0.05). Finally, patients with RCC and low TPS had a 60% lower risk of death than those with high TPS (p = 0.04, **Table 3**). These data indicate that specific intratumoral immune markers best predict patients’ risk of death independent of currently recognized risk factors, including stage, for BRCA, HNSCC, and RCC.

**Table 3.**
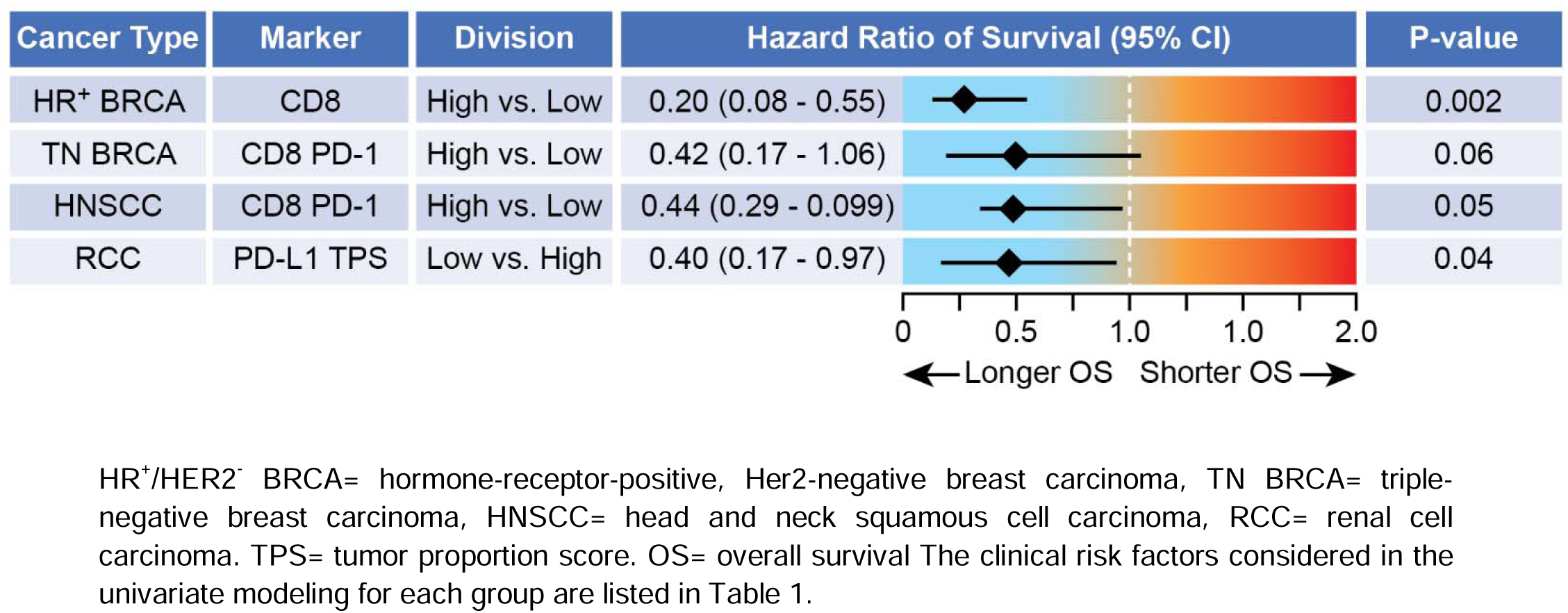
Risk of Death in Univariable Biomarker Cox Models with Clinical Risk Factor Adjustments for Cancers with Low Case Numbers.

## DISCUSSION

Expert visual interpretation of cancer tissue histomorphology on H&E stain remains the cornerstone of cancer diagnoses.^1, 2^ The results are often supplemented with select IHC staining to establish tumor cell lineage and targeted genetic testing to assess for specific oncogenic driver mutations.^4, 40^ These are critical for determining cancer classification, prognosis, and therapy. However, tissue sections contain a wide variety of additional biological information that is not routinely captured or reported. Perhaps the most critical of these is the state of the intratumoral immune cell infiltrate.

Many research studies have found that TILs and specific immune cell subsets have prognostic and predictive value for cancer patients. These include CD8^+^ and PD-1^+^ T cells, markers of active anti-tumor immunity, and FOXP3^+^ T cells, a marker of immune suppression.^39, 41–45^ Still, the results have been difficult to translate into routine practice. Diagnostic reporting for melanoma may (or may not) include the presence of a “brisk” or “non-brisk” lymphocyte response.^46^ Similarly, diagnostic reporting for colon cancer may (or may not) include a statement concerning a “Crohn’s-like” immune response.^47^ The semi-quantitative TIL scoring protocol proposed for breast cancer reporting has been validated in multiple studies. However, due to its perceived subjective scoring, it lacks broad implementation.^48^ Despite these studies and others, the only routinely performed and reported clinical assay used to assess anti-tumor immunity broadly across multiple cancer types is PD-L1 scoring, a test dependent upon pathologists’ subjective interpretation of IHC.^49^

We have addressed these deficiencies by validating and prospectively testing ImmunoProfile, a new multi-step workflow incorporating automated mIF staining, WSI capture, and ML-assisted cell quantitation to reliably capture and quantify specific immune biomarkers in FFPE tissue samples in the clinical laboratory. We found that the results were concordant with traditional IHC, reproducible over time, and yielded reportable data in >97% of the 2,023 cases tested prospectively over 3 years.

The heterogeneous nature of the specimens tested is this study’s limitation and strength. We purposefully tested all cases with sufficient tissue from all patients consented to a research protocol. Thus, there was significant variation in biopsy methods and sites, tissue types, and patients who received immune therapy, or not. As such, we can confidently state that the cohort reflects a “real world” collection of tissue specimens from cancer patients evaluated at a tertiary cancer center.

In a pan-cancer analysis, we found that high numbers of intratumoral CD8^+^, PD-1^+^, CD8^+^PD-1^+^, or FOXP3^+^ cells were associated with longer OS after controlling for significant risk factors, including cancer type, metastatic versus primary site, age, gender, and smoking. CD8+ and PD-1+ cells were independently associated with OS in a multivariable biomarker analysis. In a series of cancer-type-specific sub-analyses, we found that high numbers of CD8^+^, PD-1^+^, and/or CD8^+^PD-1^+^ cells were associated with longer OS after adjusting for risk factors, including AJCC stage, for patients with NSCLC, CRC, HR^+^/HER2-BRCA, TN BRCA, PANC, HNSCC, EGC, and OVCA despite diverse therapies. Although the substantial survival advantage that patients with high numbers of intratumoral PD-1^+^ cells have in the setting of many cancer types might be unexpected, our data suggest that PD-1, a marker of T-cell activation as well as exhaustion, may better serve as a marker of effective anti-tumor immunity than CD8 alone for some cancer types.^50^

In contrast, we found that patients with BLAD and high numbers of FOXP3^+^ cells had longer OS. We also found that patients with RCC and *low* PD-L1 TPS scores (below the group median) had longer OS. As these results have been found in prior retrospective studies, they likely reflect a unique immunobiology inherent to these tumors.^51, 52^ Greater clarity concerning these results may come with more detailed analyses.

In summary, the quantitative evaluation of defined intratumoral immune cell populations, most critically CD8^+^, PD-1^+^, and CD8^+^PD-1^+^ cells, provides prognostic information not captured by the current clinical staging and risk assessment systems across many cancer types. In this way, ImmunoProfile both validates and extends the clinical validation of ImmunoScore, a semi-automated staining, imaging, and scoring platform that has undergone rigorous clinical testing and improves the stratification of patients with colon cancer better than the AJCC TNM staging system.^22, 24^ With recent advances in automated staining, imaging, and scoring technologies, we can anticipate that additional, highly specified lymphoid subsets identified in research studies^53^ will be reliably captured and reported in routine clinical practice.

## Supporting information

Alessi_etal_SupplTable1

Alessi_etal_SupplMethods

Alessi_etal_SupplFiguresTables

## Data Availability

All data produced in the present work are contained in the manuscript.

## ACKNOWLEDGEMENTS

We thank the Dana-Farber Cancer Institute and its patients and donors, whose charitable contributions supported this study. We would also like to thank the following individuals who supported this effort: Bill Hahn, Laurie Glimcher, Jeffrey Golden, Cathleen Quade, Athena Petrides, Jane Song, Eric Reed, and Shadan Shafieha. The IRB of the Dana-Farber Cancer Institute gave ethical approval for this work. This project was conducted under DFCI protocols 11-104, 17-000, 20-000, and 22-176.

## CONFLICTS OF INTEREST

SR receives research support from Bristol Myers Squibb, KITE/Gilead, and Surface Oncology. SR is an SAB member for Immunitas Therapeutics. LMS receives research support from Bristol Myers Squibb and Genentech and has received consulting income from Genentech, Lilly, and AstraZeneca. PKS is a member of the BOD of Glencoe Software and SAB for RareCyte, NanoString, and Montai Health; he consults for Merck and Novartis.

## Notes

### Competing Interest Statement

The authors have declared no competing interest.

### Funding Statement

This study was funded by the Dana-Farber Cancer Institute and its charitable contributors.

### Author Declarations

The IRB of the Dana-Farber Cancer Institute gave ethical approval for this work.

