## Supplementary material for "Clinical Immunoprofiling Reveals that High Numbers of CD8^+^ and PD-1^+^ Cells Predict Superior Patient Survival Across Major Cancer Types Independent of Major Risk Factors": Alessi_etal_SupplMethods

**Alessi et al., SUPPLEMENTARY METHODS**

***Patient Consent, Clinical Laboratory, and Clinical Materials***

All patients consented to institutional a research use protocol for this study. A pathologist reviewed Hematoxylin and eosin (H&E) stained slides for all specimens to confirm tumor content before specimen staining. All validation studies, patient tissue staining, and analyses were performed in a CLIA-certified laboratory (License #22D2156930) in the Department of Pathology at Brigham & Women’s Hospital. Key novel features developed for the ImmunoProfile assay included: 1) automated test ordering and consent verification within the electronic medical record, 2) a web-based digital microscopy environment for WSI review, 3) an interactive web-based workflow to annotate and approve tumor-stromal borders and regions of interest (ROIs), 4) integrated software tools for ML-assisted automated cell phenotyping, scoring, and results reporting, and 5) a HIPAA-compliant data warehouse for storing test results and digital images.

***Immunostaining Optimization***

We first optimized single-plex chromogenic immunohistochemistry (cIHC) using a Bond Rx autostainer and tonsil tissues under various staining conditions. The results were reviewed, and the optimal conditions were selected by a pathologist (SR). The conditions were adapted to single-plex immunofluorescence (sIF) stains performed on a Bond Rx autostainer, the results of which were reviewed and approved by a pathologist. Finally, the protocols for each sIF were combined into a multiplex immunofluorescence (mIF) protocol performed on the Bond Rx following previously published protocols and approved by a pathologist.^1–3^ We then verified the optimized conditions by staining 10 FFPE samples of representative tumor types (bladder, head and neck, and lung cancer) and additional normal tonsils by cIHC, sIF, and mIF. The pathologist approved all conditions after a qualitative and quantitative review of the cases. The clinical validation documents, including IHC vs. MIF comparability studies, inter-operator reproducibility studies, inter-staining reproducibility studies, and others, were approved by the CLIA laboratory director (NL).^4,5^

***ImmunoProfile Staining Protocol***

The final mIF panel consisted of 5 markers- CD8, FOXP3, PD-1, PD-L1, and a tumor marker.^6–8^ The tumor marker was cytokeratin (AE1/AE3) for epithelial cancers, PAX8 for renal cell carcinomas, and SOX10 for melanomas. DAPI is used as a counter stain to detect nuclei. MIF was performed on the BOND RX fully automated stainer
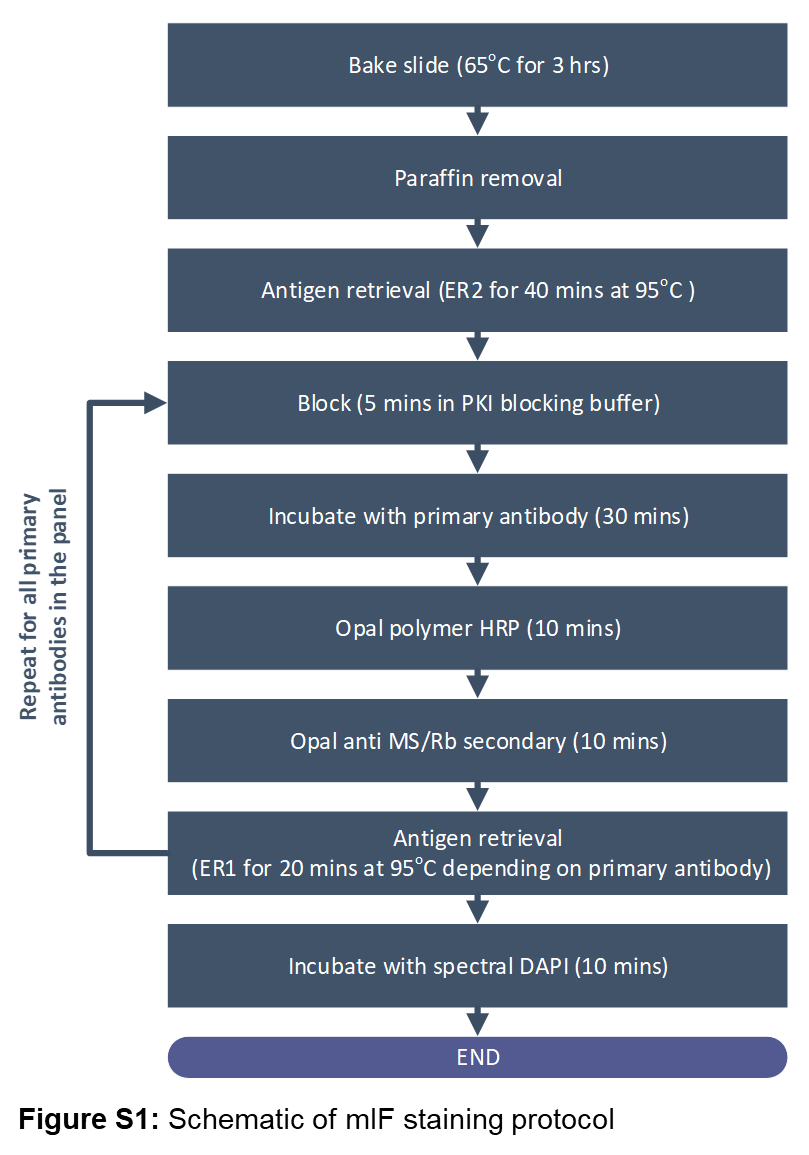
(Leica Biosystems, Danvers, MA). The workflow for mIF staining is shown in **Figure S1**.^2,9,10^ To summarize, FFPE blocks were sectioned onto positively charged glass microscopic slides in 5-μm thick sections before staining. Before loading into the BOND RX, slides were baked for 3 hours at 65°C. The BOND RX pre-programmed and executed the deparaffinization, rehydration, and antigen retrieval. Next, slides were serially stained with primary antibodies, starting with anti-FOXP3. The incubation time was 30 minutes per primary antibody. Subsequently, anti-mouse plus anti-rabbit Opal Polymer Horseradish Peroxidase (Opal Polymer HRP Ms + Rb, Akoya Biosciences, Marlborough, MA, Cat. ARH1001EA) was applied as a secondary label with a 10-minute incubation time. The process was repeated starting from the antigen retrieval step for each additional antibody in the following order: FOXP3, PD-L1 (Programmed death ligand 1), cytokeratin, PD-1, and CD8. The signal for antibody complexes was labeled and visualized by their corresponding Opal Fluorophore Reagents (Akoya) by incubating the slides for 10 minutes. Finally, slides were stained with DAPI for 10 minutes, washed in deionized water, air dried, mounted with Prolong Diamond Antifade Mountant (Life Technologies, Carlsbad, CA, Cat. P36965), and stored in light-proof boxes at 4°C. The target antigens, antibody clones, dilutions for markers, and antigen retrieval details are listed in **Table S1**.

**Table S1. *Target antigens, antibody clones, antibody dilutions, and antigen retrieval conditions***

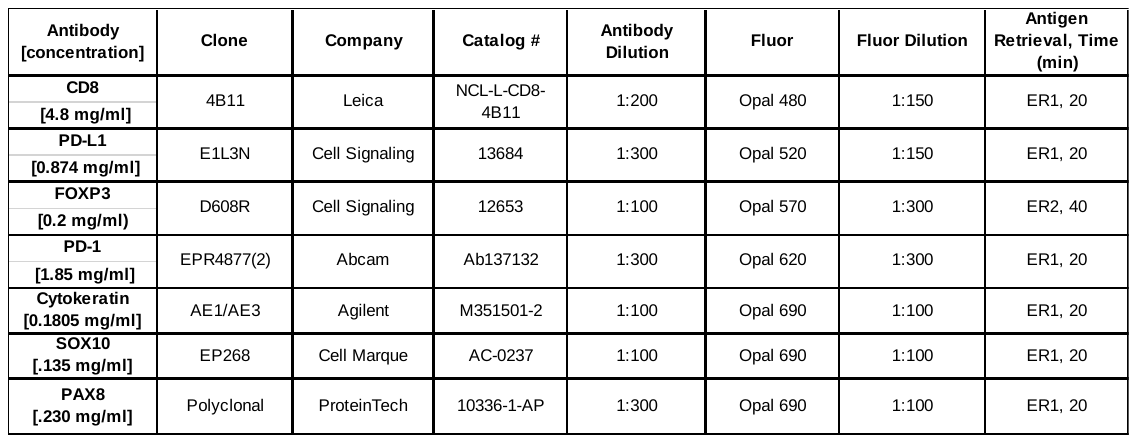

***Clinical Controls and PD-L1 Control Tissue Microarray (TMA)***

The IP cohort study began in November 2018. Qualitative tonsils and tumor tissue were used as controls from November 2018 to December 2020. At the end of 2020, a tissue microarray was created using cores from cancer tissues known to be positive and negative for PD-L1 as determined by the ImmunoProfile results from cases analyzed.^11,12^ The TMA was used as a quantitative control in addition to the qualitative tonsil control for all IP cases done from January 2021. Each core (0.6 mm in diameter) fit into one ROI used for analysis (one 20X field of view correlates to 925 µm x 693 µm). Regions for use in the TMA were annotated by a scientist on H&E slides and approved by a pathologist.

The array (named *PD-L1 control TMA*) contained three cores from a PD-L1 positive colorectal cancer case, which was used as the PD-L1 positive control. These cores also expressed all other markers in the panel to a relatively high degree. The TMA also contained three cores from a PD-L1 negative bladder cancer case, used as the PD-L1 negative control. The PD-L1 negative cores also had relatively low expression of other markers (except DAPI and cytokeratin) in the panel.

The results from staining and analysis of *PD-L1 control TMA* were used to test intra-run reproducibility for staining, inter-run reproducibility for staining, and inter-run reproducibility for scanning (see *Supplementary Figure 2*).

***ImmunoProfile Sign-out Tool***

To facilitate workflow through the image analysis pipeline, an in-house custom interface for scientists and pathologists to collaborate for ROI approvals, analysis review, and final report generation called the *ImmunoProfile Sign-out Tool* was created. This tool interfaces with hospital-wide databases to import newly ordered ImmunoProfile cases and pull relevant patient information needed to review the case. It supports qualitative and quantitative staining control approvals, region of interest (ROI) annotations, tumor-stromal interface annotations, analysis output generated by the scientists, and built-in algorithms to compute cell density measurements and PD-L1 scores from the output data. Furthermore, after the signing out of a case, this system also interfaces with other databases to send and display results for researchers and clinical investigators.

***Clinical Workflow***

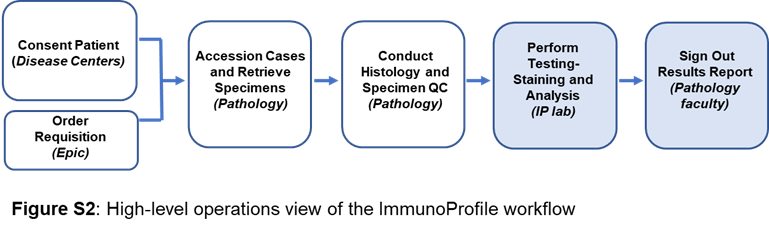
 A custom-built *Electronic Consent Tool* confirmed patient consent to one of two research protocols stored in a research database (OncDRS, DFCI, **Figure S2**). FFPE biopsy materials from consented patients were obtained from the clinical laboratory, pathologists reviewed subsequent H&Es to confirm tumor content, and slides were stained using the automated protocols (**Figure S2**). An important challenge to the clinical workflow was to ensure tissue availability and sufficient tumor content for IP testing. FFPE material derived from excisional or core needle biopsies with at least 100 tumor cells was required. The most common reason for failure to proceed with patient testing was insufficient tumor cells in the biopsy sample, as seen for NGS clinical testing^13^.

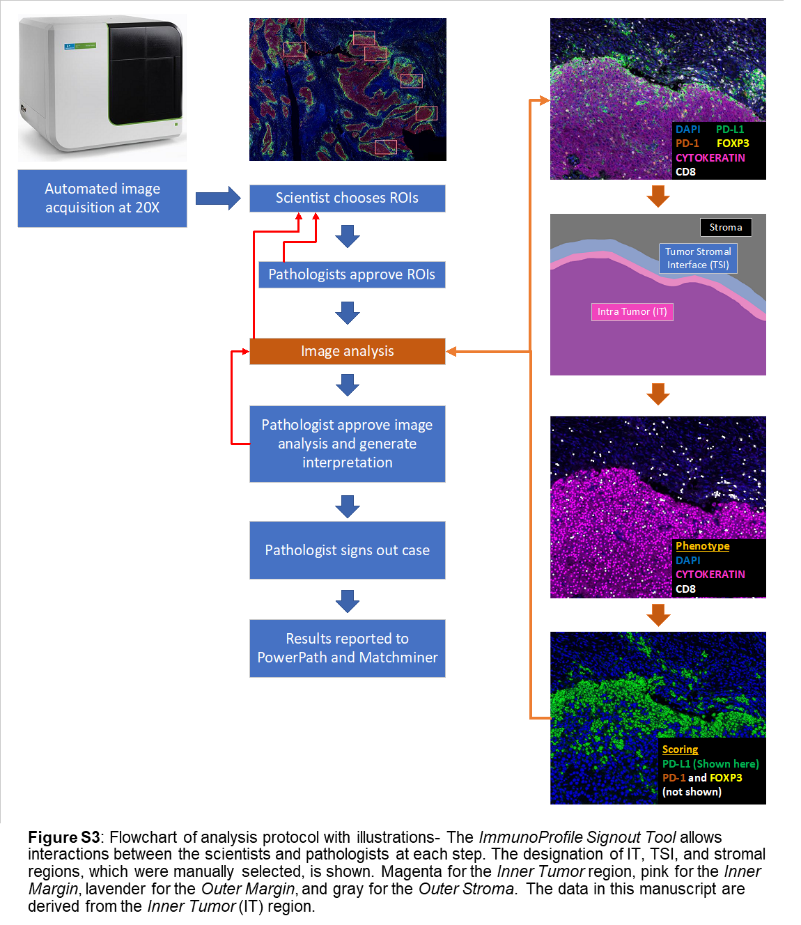
 Whole slide images were then acquired at 20x resolution using Akoya Bioscience’s PhenoImager MOTiF™ imaging platform. Subsequent steps are outlined below (**Figure S3**).^1^ First, regions of interest (ROIs) representative of the overall tumor microenvironment were chosen by a scientist and approved by a pathologist. Intra-Tumor (IT) and Tumor Stroma Interface (TSI) regions were then annotated by the scientist with an internally developed GNU Image Manipulation Program (GIMP, https://github.com/jason-weirather/pythologist). The ROIs were spectrally unmixed and analyzed using inForm 2.4.8 (Akoya). Analysis within inForm involved cell segmentation using a nuclear marker (DAPI) and additional membranous markers such as cytokeratin and/or CD8. Tumor and cytotoxic T-cells were phenotyped based on the expression of cytokeratin and CD8, respectively. Other (non-tumor and non-cytotoxic T cells) were phenotyped based on the lack of expression of cytokeratin and CD8. Identification of FOXP3, PD-1, and PD-L1 was achieved by setting an intensity threshold individually for these markers by the scientists analyzing the case based on the scientist’s judgment of staining patterns and intensity.

Scientists uploaded all images and annotations into the ImmunoProfile Sign-out Tool (**Figure S3**), which was reviewed by a pathologist and approved at every step. The custom-developed algorithmic capabilities were used to automatically calculate densities of CD8+, PD-1+, CD8+/PD-1+, and FOXP3+ populations for IT, TSI, and IT+TSI (total) regions when applicable (<https://github.com/jason-weirather/pythologist>).^9,10^ PD-L1 scores, including TPS (Tumor Proportion Score), CPS (Combined Positive Score), IPS (Immune Proportion Score), and IC (Immune Cell Area Score), were also automatically computed (**Figure S3**). The results were reviewed by pathologists, approved, and finalized within the ImmunoProfile Signout Tool.

**ImmunoProfile Success and Failure Rates**

2,537 cases were processed and available in the IP database when the test was concluded. 119 were signed out as failed cases for any reason, representing 4.6%. The most frequent reasons for failure were insufficient tumor material (34%), poor DAPI staining (12%), poor staining on multiple IF markers (11%), the cancer type not supported by the assay (9%), and not further specified (20%). A few failures were attributed to poor cell segmentation, tumor staining, high background, and inadequate sample handling (14%). We could not obtain complete clinical information for all patients, so the number of patients with clinical follow-up data was fewer than the number tested.

**Statistical Analysis**

Overall survival (OS) was defined as the time interval between the date of biopsy and death from any cause. The follow-up of patients alive at the time of data retrieval was censored at the date of last contact. For cancer subtypes with more than 100 cases and at least 50 deaths, immune cell densities were grouped according to tertiles of their respective distributions; otherwise, median splits were used. CPS and TPS were also evaluated using pre-specified, disease-specific cut points.

Statistical relationships between the biomarkers and OS were evaluated for the entire pan-cancer sample and separately for each cancer type. The distributions of OS were estimated using the Kaplan-Meier method and compared using log-rank testing; 95% confidence intervals were calculated using log(-log) methodology. Median follow-up was based on Kaplan-Meier estimation with an inverted censor.

The associations between each immune cell density measurement and OS were assessed using Cox proportional hazards models adjusted for known clinical risk factors, such as American Joint Committee on Cancer (AJCC) stage, microsatellite instability (MSI) status (colorectal cancer), age, alcohol(current/former/never/unknown), and tobacco use (current/former/never/unknown), and sex. The pan-cancer model was additionally stratified by cancer type. For each cancer, risk factors were identified, prioritized, and selected before analysis and were based on clinical practice and known relationships with survival. To avoid overfitting, an appropriate number of prioritized risk factors was included in each model according to the cohort size and number of deaths.

Multivariable models were fit for any cancer type with at least 30 deaths. Immune cells with adjusted Wald p-values ≤ 0.1 from the single marker analyses were carried forward as candidate biomarkers in the multivariable setting. Iterative model building was then performed in which candidate biomarkers were added one at a time to a risk factor-adjusted model using likelihood-ratio testing. Biomarker inclusion stopped when no additional candidate biomarker resulted in a statistically significant chi-squared p-value. Hazard ratios are presented with 95% confidence intervals. Model fitting was performed using SAS 9.4 (SAS Institute Inc., Cary, NC, USA).

**Data Availability**

The inability to access control standards, especially immune markers, has impeded the acceptance and implementation of quantitative tissue biomarker assays across clinical laboratories. One solution is to provide a web-based data warehouse containing a set of stained control tissues that have been imaged, annotated and scored using the workflow deployed for this study. We analyzed consecutive FFPE tonsil sections that were stained, imaged, annotated, and scored over 3.25 years using ImmunoProfile. These data are available as controls for developing and validating institution-specific workflows and quality control comparisons (*Supplementary Materials*). In addition, the entire set of images and quantitative data derived from the control cancer tissue microarray (control TMA) and the patient samples are available for reference against institution-specific datasets.
