## Supplementary material for "Clinical Immunoprofiling Reveals that High Numbers of CD8^+^ and PD-1^+^ Cells Predict Superior Patient Survival Across Major Cancer Types Independent of Major Risk Factors": Alessi_etal_SupplFiguresTables

**Alessi et al., Supplementary Figure 1.**

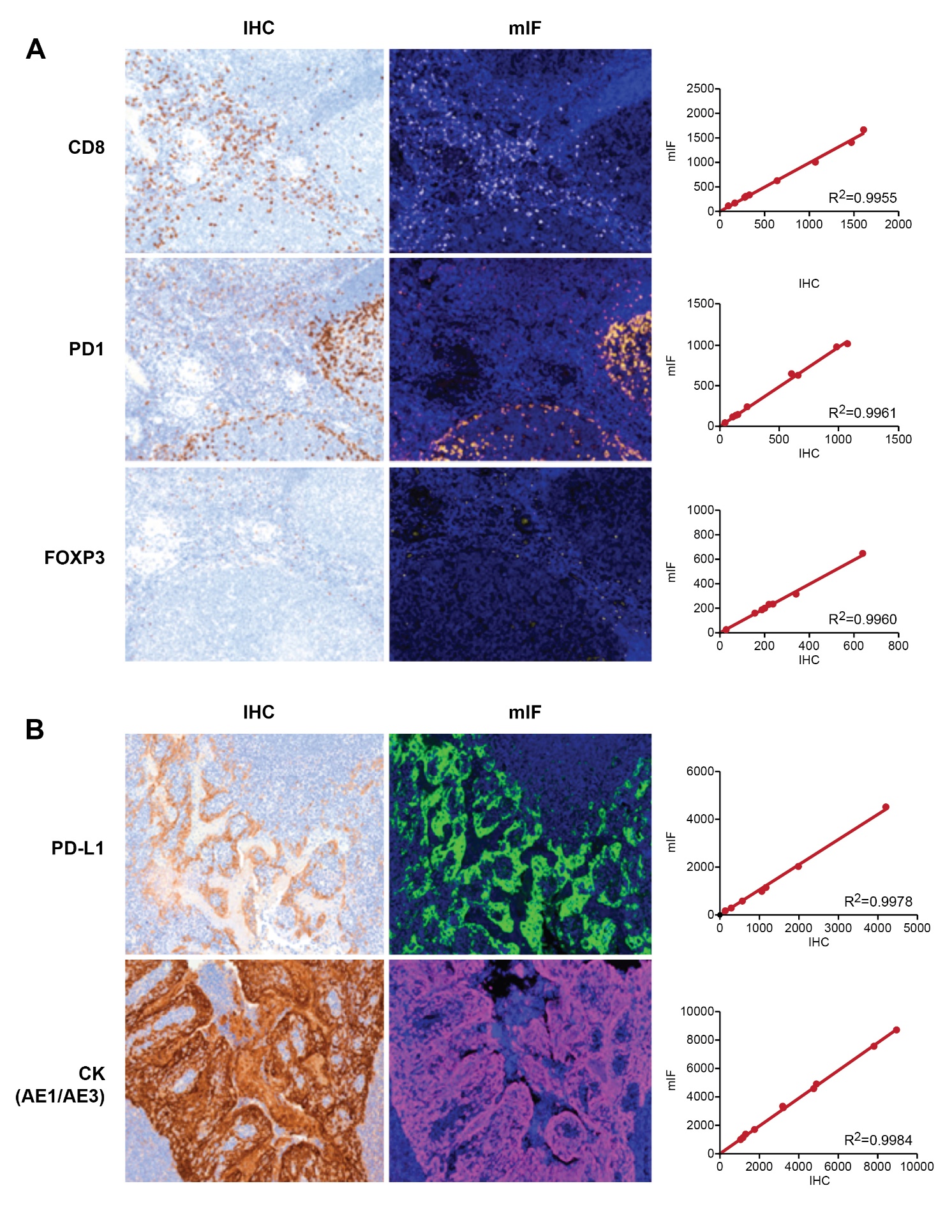

**Supplementary Figure 1.** Select data from the ImmunoProfile validation studies performed within the CLIA-certified clinical laboratory. **A:** Example images from chromogenic immunohistochemical stained (IHC*, left panels*) and the corresponding single-channel immunofluorescence from multiplex-stained (mIF, *middle panels*) serial sections with antibodies targeting the indicated immune cell biomarkers on formalin-fixed paraffin-embedded (FFPE) tonsillar tissue. The number of positive staining cells detected by IHC and mIF for the indicated markers across control tonsil tissues and their corresponding correlation coefficients (R^2^). **B:** Example images from single chromogenic immunohistochemical stained (IHC*, left panels*) and the corresponding single-channel immunofluorescence from multiplex-stained (mIF, *middle panels*) serial sections with antibodies targeting the indicated biomarkers on formalin-fixed paraffin-embedded (FFPE) tonsillar tissue. The number of positive staining cells detected by IHC and mIF for the indicated markers across control tonsil tissues and their corresponding correlation coefficients (R^2^).

**Supplementary Figure 2.**

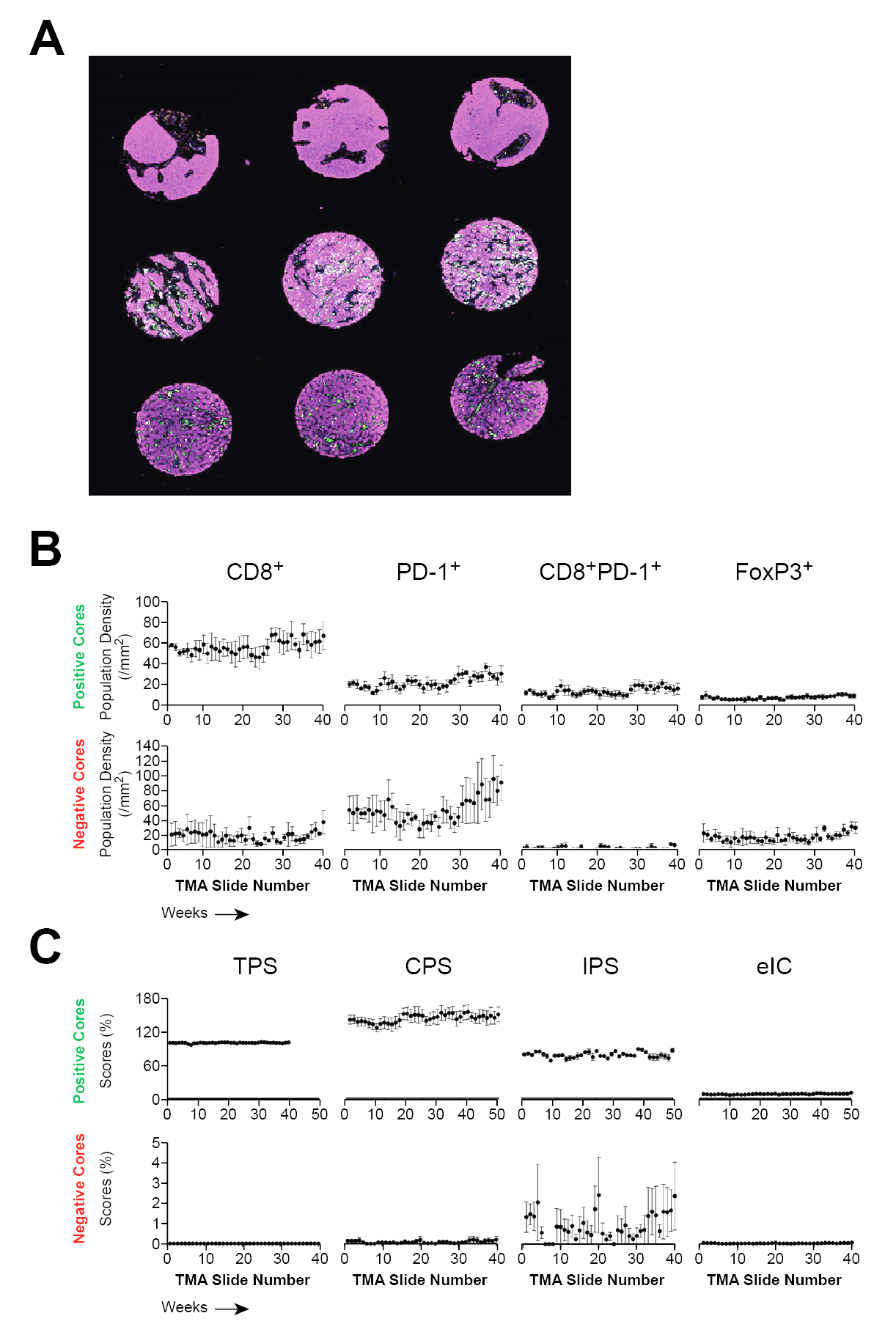

**Supplementary Figure 2.** IP reproducibility over time. **A:** MIF image of a control tissue microarray (TMA) stained during a routine patient sample staining run and showing positive staining of cancer epithelial cells (cytokeratin [CK] stain, *pink*), PD-L1 (*green*), and CD8 (*white*) and imaged at 200x original magnification. Note that the control TMA includes cancers that are immunologically “cold” (*top row*) and “hot” (*middle and bottom rows*). **B:** The control TMA was stained, imaged, and quantified weekly with patient samples. Quantitation of the cell densities (cells per mm^2^) for the indicated immune cell markers in the “hot” (*positive cores*) and “cold” (*negative cores*) tumors in serial sections of the control TMA stained and quantified serially over the indicated number of weeks is shown. **C:** The control TMA was stained, imaged, and quantified weekly with patient samples. Quantitation of PD-L1 tumor proportion scores (TPS), combined proportion scores (CPS), immune proportion scores (IPS), and estimated immune cell scores (eIC) in the “hot” (*positive cores*) and “cold” (*negative cores*) tumors in serial sections of the control TMA stained and quantified serially over the indicated number of weeks is shown.

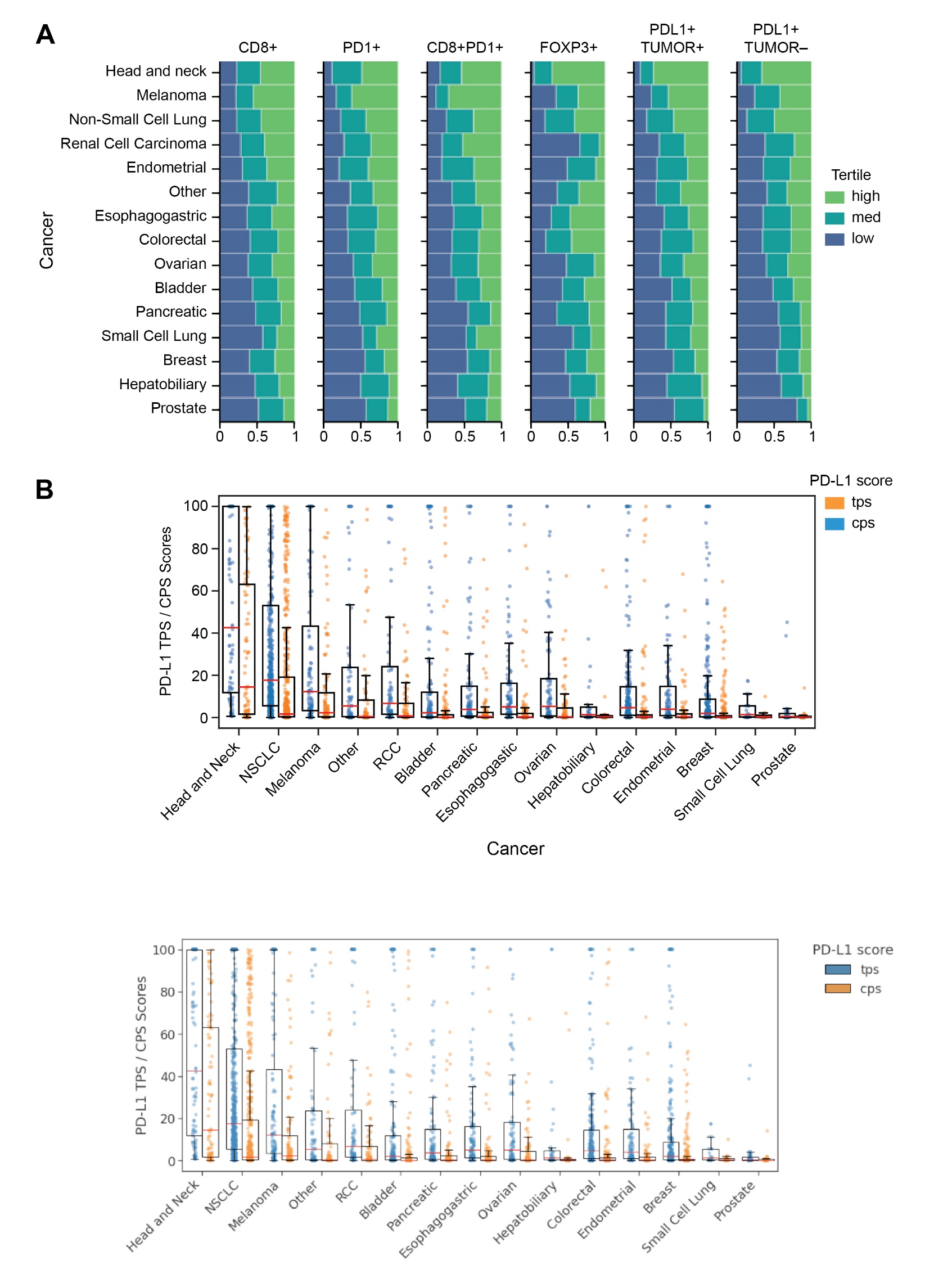
**Supplementary Figure 3.**

**Supplementary Figure 3.** The distribution of immune biomarker scores by cancer types. **A:** The relative proportion of patients of each cancer type with biomarker cell densities that are high, medium, or low by tertile divisions for the pan-cancer cohort. Note that each cancer type has a proportion of patients with tumors in the highest and lowest tertiles for each of the biomarkers. **B:** The distribution of PD-L1 tumor proportion scores (tps) and combined proportion scores (cps) across cases divided by cancer types. The median (*orange line*), 25^th^ and 75^th^ centile (*boxes*), and extremes (*brackets*) are indicated. Note that each cancer type has a proportion of patients with tumors that have very high or low TPS and CPS scores.

**Supplementary Figure 4.**

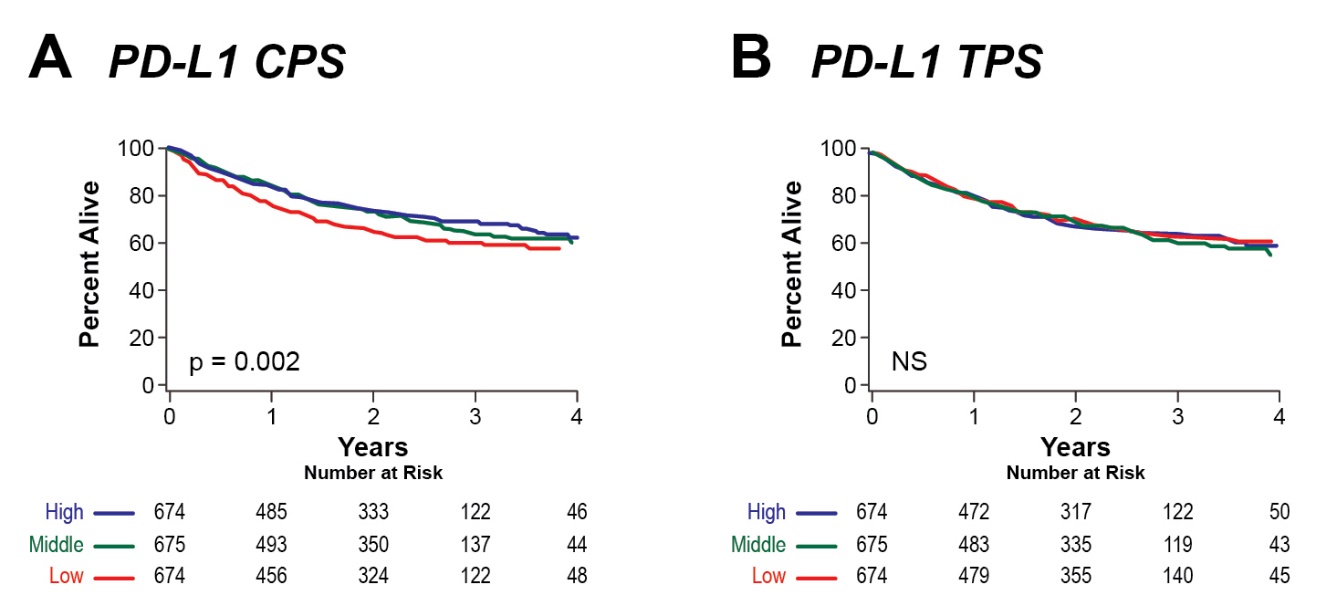

**Supplementary Figure 4.** Kaplan-Meier curves of overall survival (OS) of patients in the pan-cancer cohort divided according to their PD-L1 combined proportion scores (CPS) and PD-L1 tumor proportion scores (TPS) into high (blue), middle (green), and low (red) tertiles based on their respective biomarker distributions. The number of patients at risk at the indicated number of years from the biopsy is indicated. Significant p-values by the log-rank test are shown. NS = not significant.

**Supplementary Figure 5.**

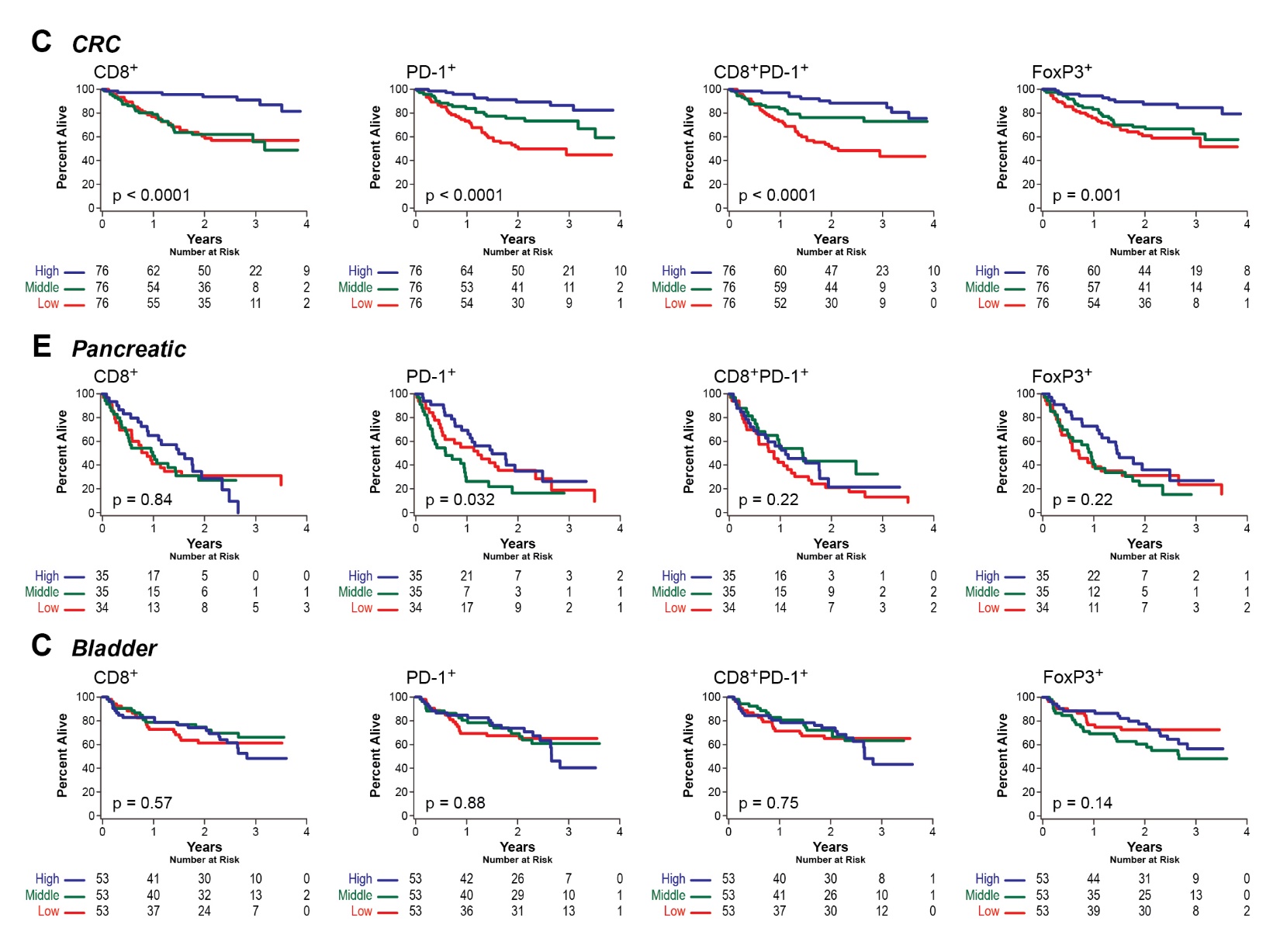

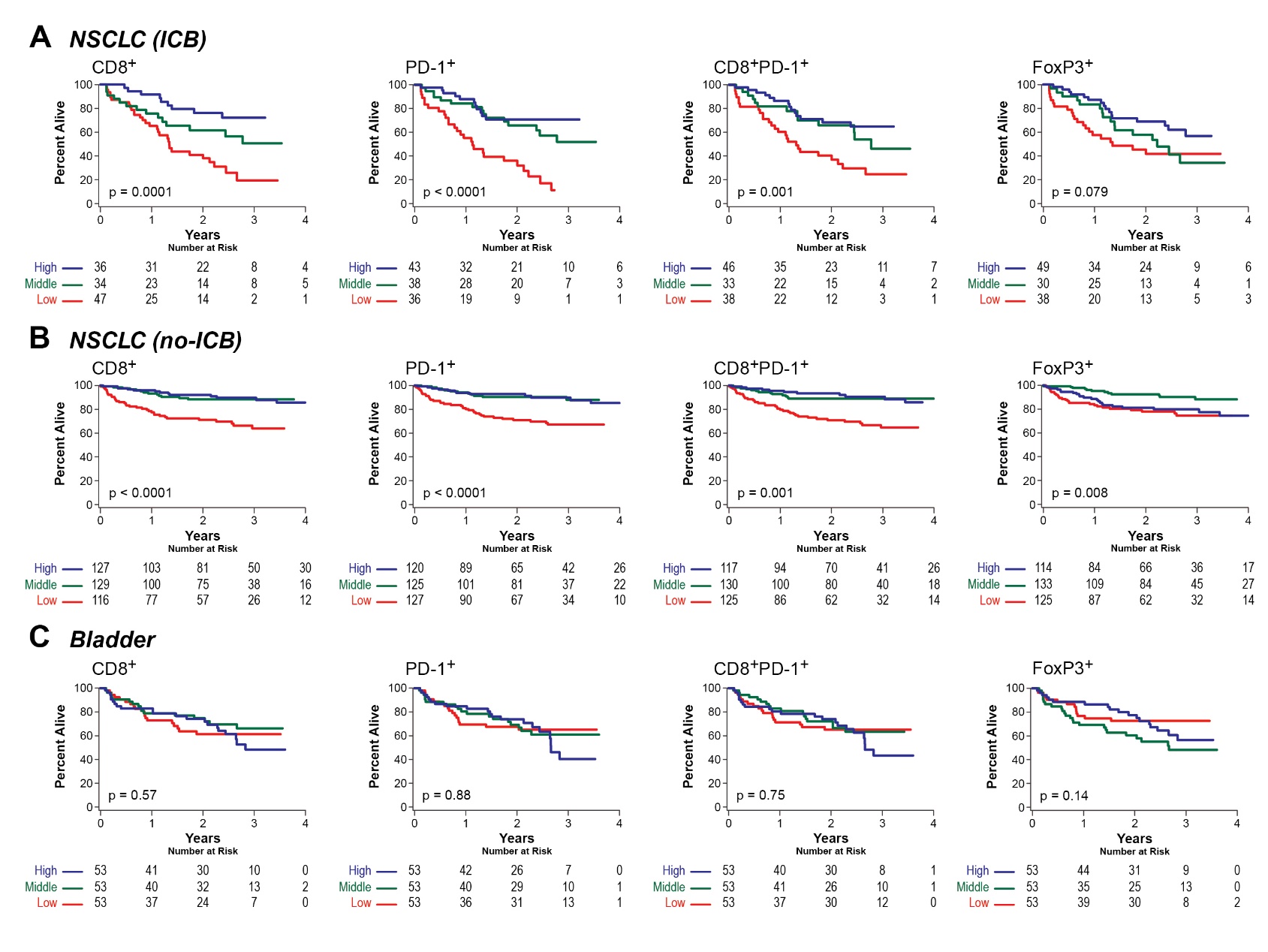

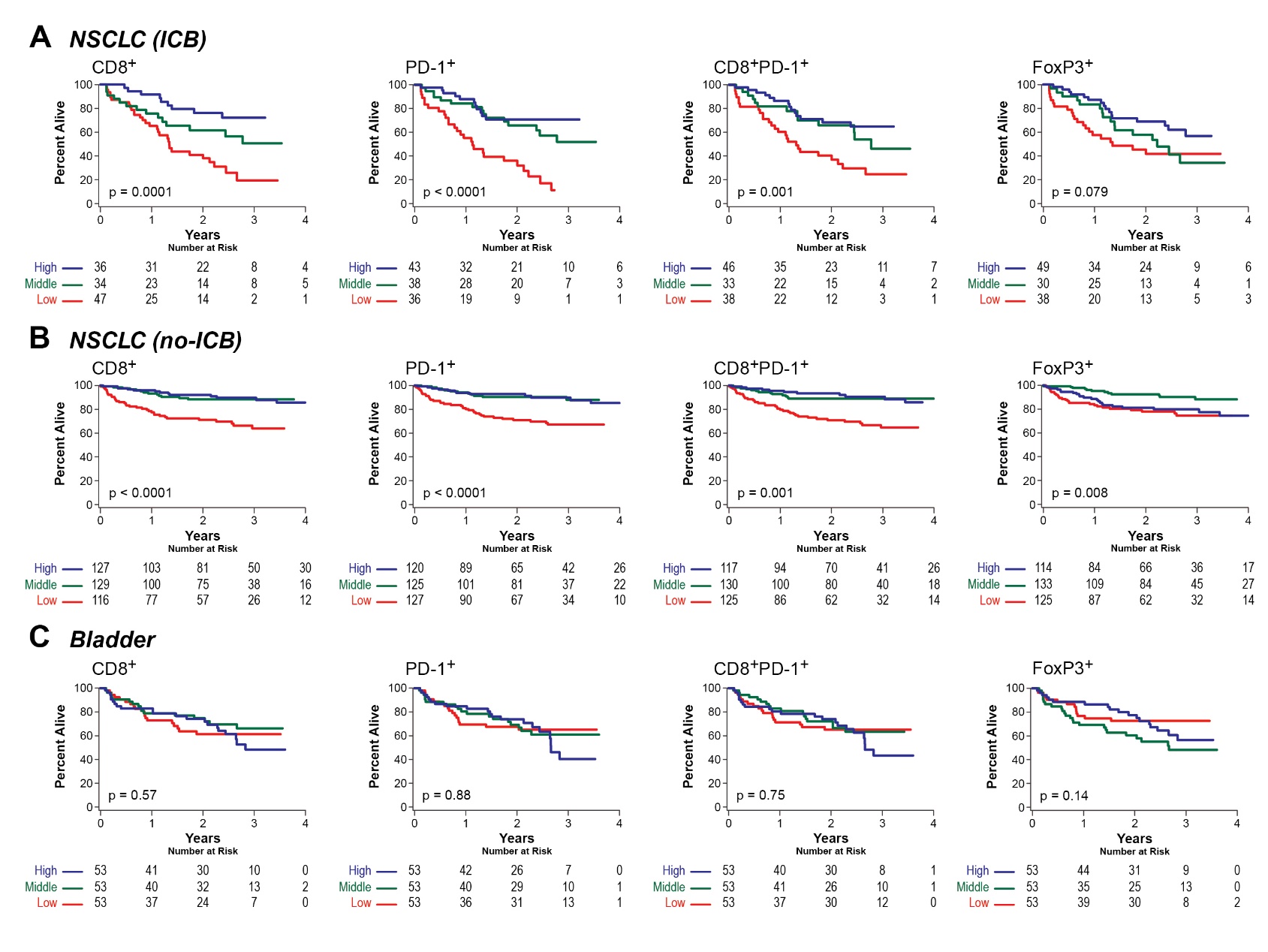

**Supplementary Figure 5.** Kaplan-Meier curves of overall survival (OS) according to tertiles of the biomarker distributions. The patients were divided according to their CD8^+^, PD-1^+^, CD8^+^PD-1^+^, and FoxP3^+^ immune cell densities into high (blue), middle (green), and low (red) tertiles for each respective group. The number of patients at risk at the indicated number of years from diagnosis is shown for **A:** Patients with pancreatic cancer (PAN, n = 104), **B:** Patients with bladder cancer (BLAD, n = 159). The p-values by the log-rank test are indicated.

**Supplementary Figure 6.**

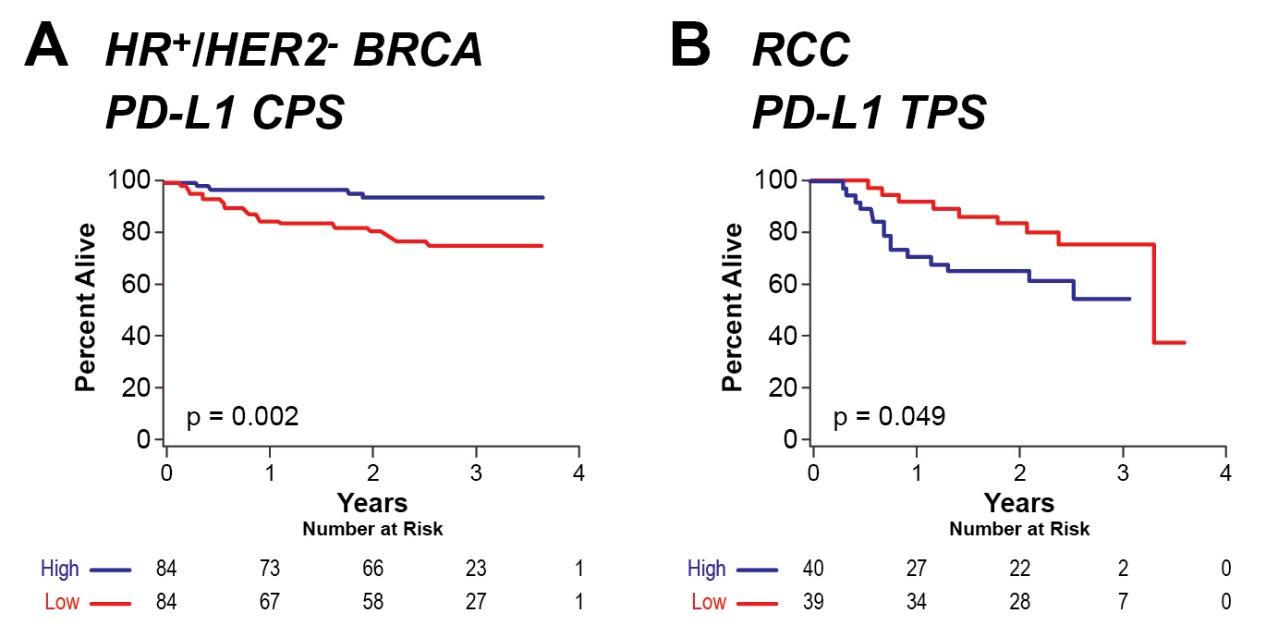

**Supplementary Figure 5.** Kaplan-Meier curves of overall survival (OS) for biomarkers divided at the median. **A:** HR^+^/HER2^-^ BRCA patients were divided at the median according to their PD-L1 CPS scores into high (blue) and low (red) groups. **B:** RCC patients were divided at the median according to their PD-L1 TPS scores into high (blue) and low (red) groups. The number of patients at risk at the indicated number of years from the biopsy is indicated. The p-values by the log-rank test are indicated.

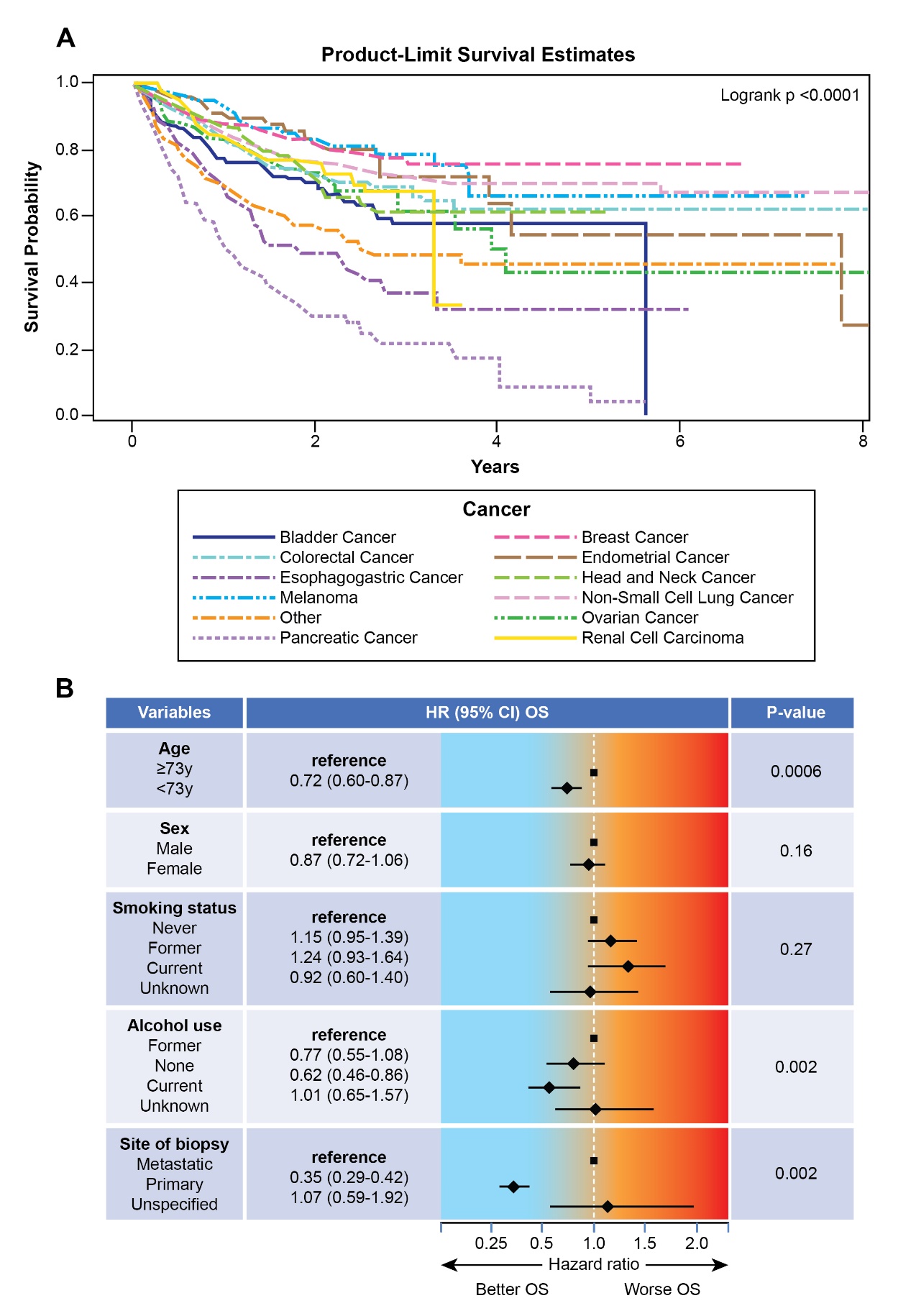
**Supplementary Figure 7.**

**Supplementary Figure 7.** Risk factor characteristics for the pan-cancer cohort. **A:** Patient overall survival (OS) over time (in years) divided according to major cancer type (log-rank p < 0.0001). **B:** Patients’ risk of death according to the indicated risk factor status.

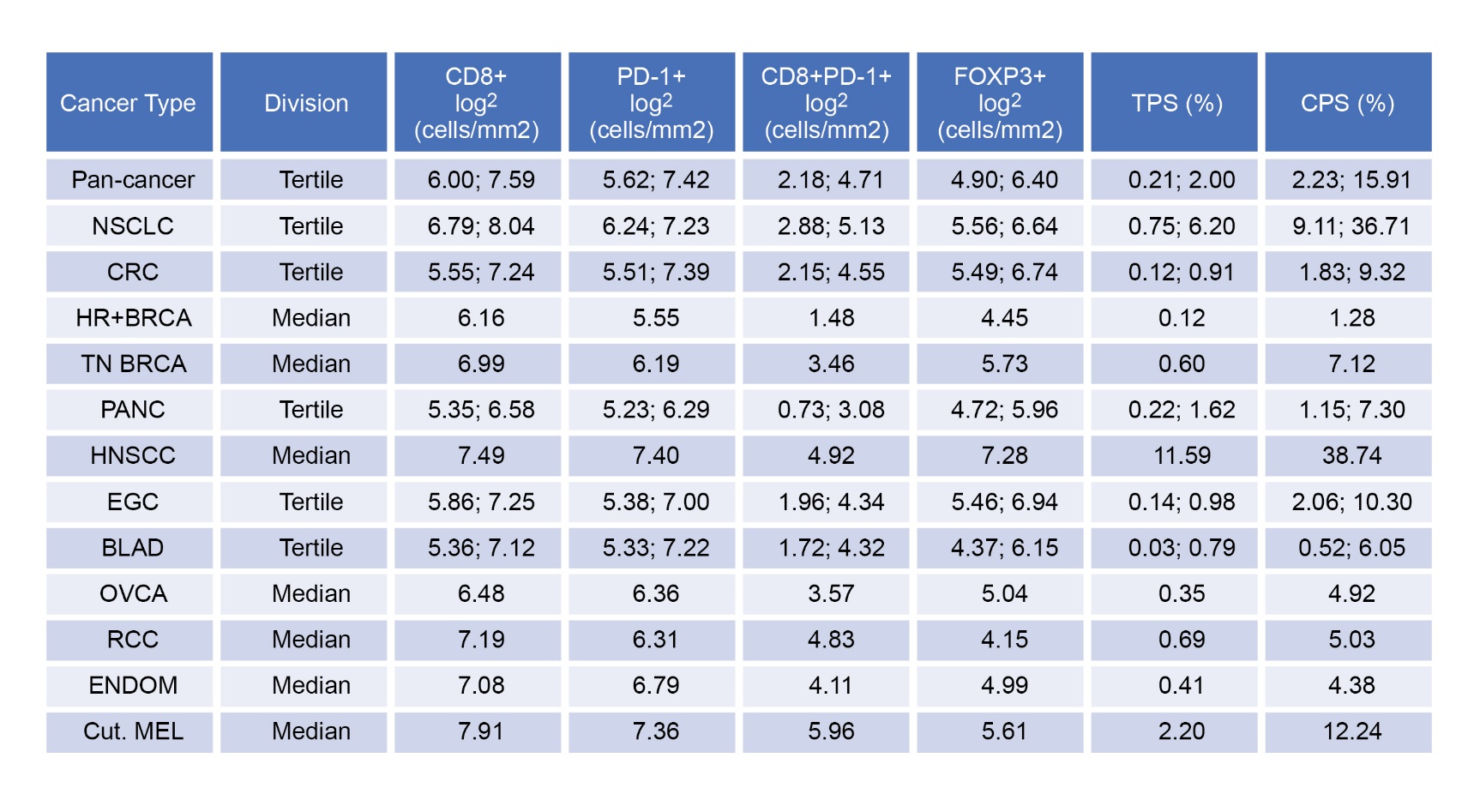
**Supplementary Table 2. Raw tertile and median division values**

NSCLC= non-small cell lung cancer, CRC= colorectal carcinoma, HR^+^/ Her2^-^ BRCA= hormone-receptor-positive breast carcinoma, TN BRCA= triple-negative breast carcinoma, PANC= pancreatic carcinoma, HNSCC= head and neck squamous cell carcinoma, EGC= esophagogastric carcinoma, BLAD= bladder carcinoma, OVCA= ovarian carcinoma, RCC= renal cell carcinoma, ENDO= endometrial carcinoma; cut. MEL= cutaneous melanoma.

**Supplementary Table 3. PD-L1 univariable analysis with risk factor adjustments**

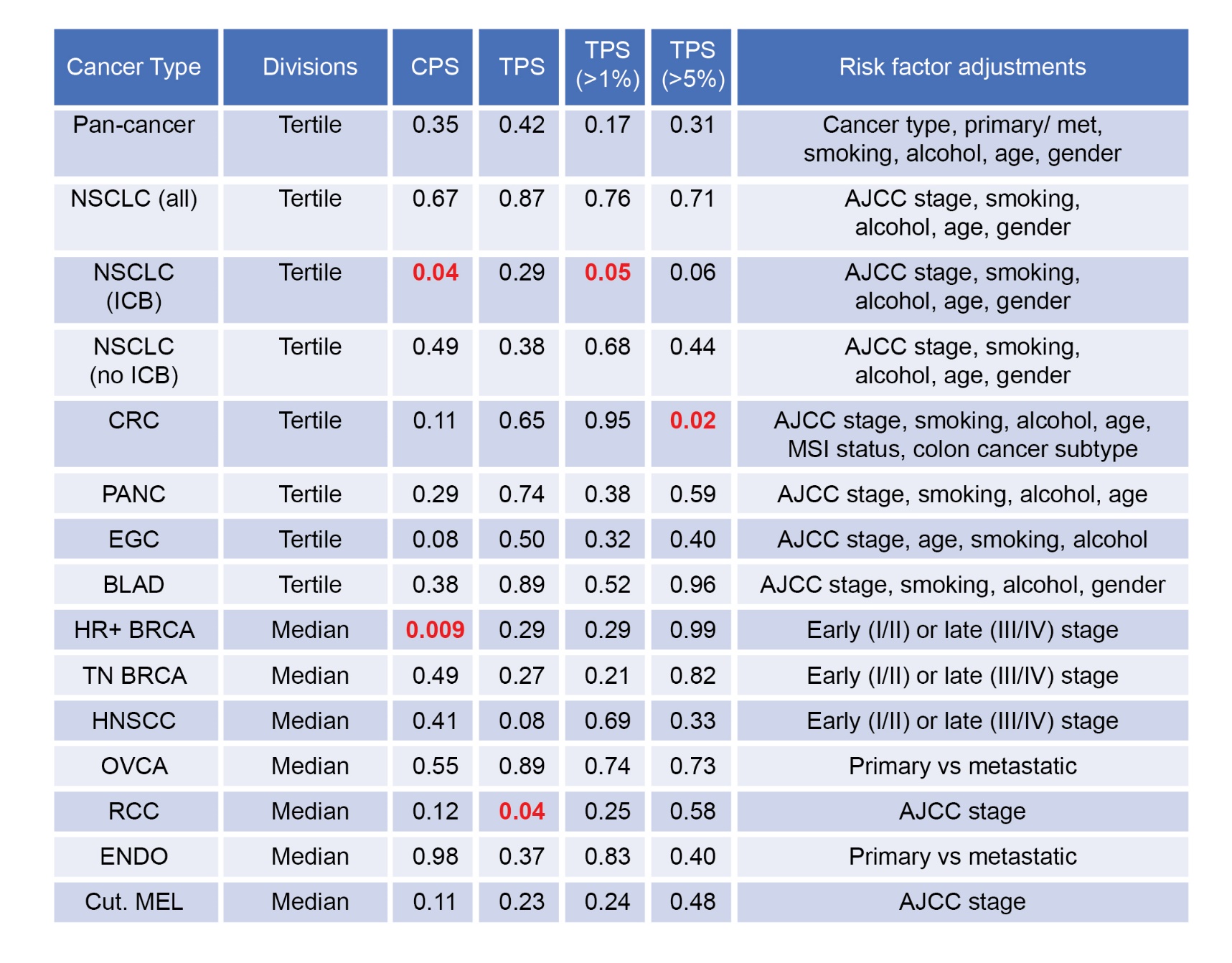

NSCLC= non-small cell lung cancer, no-ICB= no treatment with immune checkpoint blockade, ICB= treatment with checkpoint blockade, CRC= colorectal carcinoma, PANC= pancreatic carcinoma, EGC= esophagogastric carcinoma, BLAD= bladder carcinoma, HR^+^ BRCA= hormone-receptor-positive breast carcinoma, TN BRCA= triple-negative breast carcinoma, HNSCC= head and neck squamous cell carcinoma, OVCA= ovarian carcinoma, RCC= renal cell carcinoma, ENDO= endometrial carcinoma; cut. MEL= cutaneous melanoma. CPS= combined proportion score, TPS= tumor proportion score, AJCC= American Joint Committee on Cancer. Significant p values (≥ 0.05) are indicated in red.

**Supplementary Figure 8.**

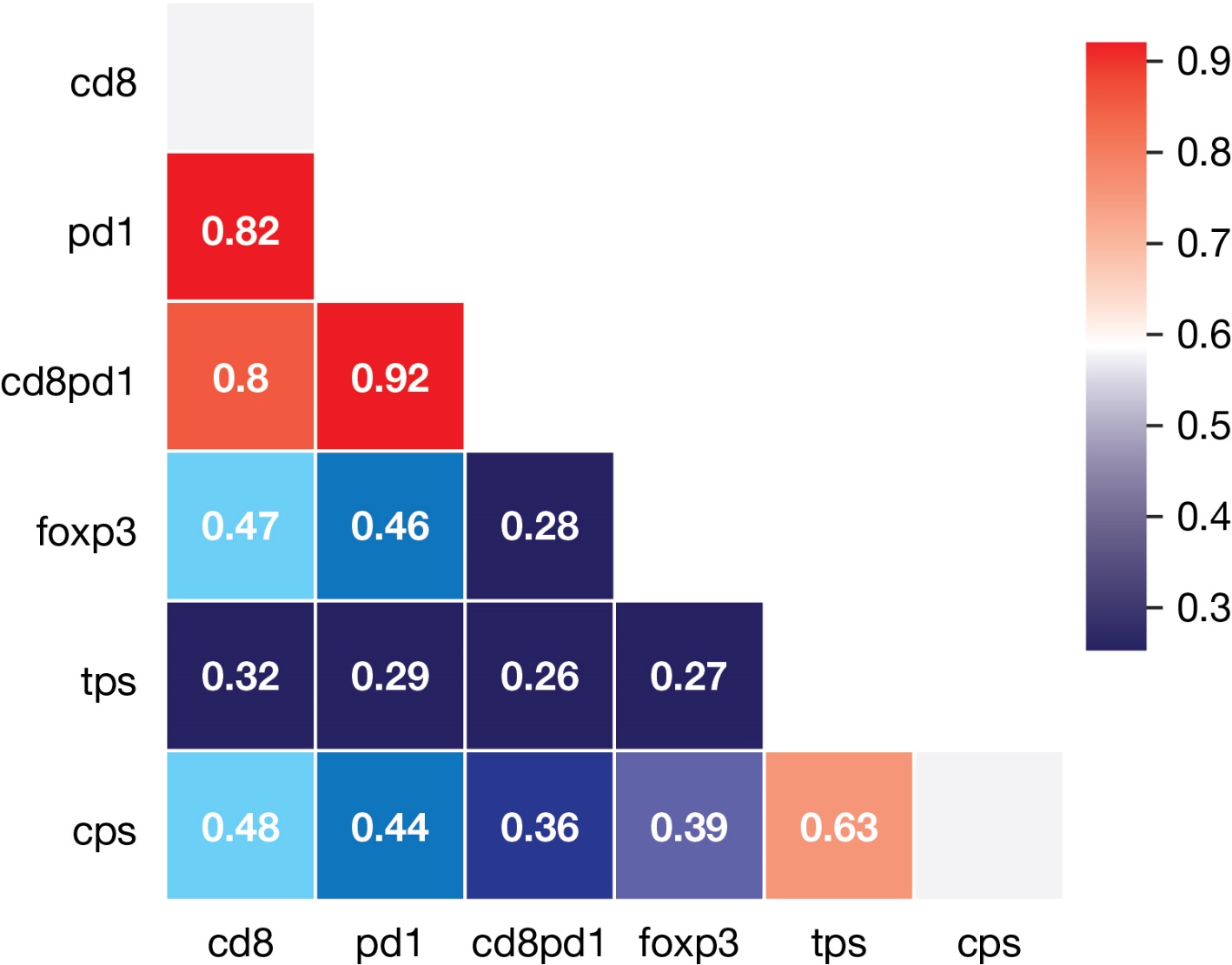

**Supplementary Figure 8.** Correlation coefficients between immunological biomarkers. The correlation coefficients between the indicated tissue biomarkers (cells per mm^2^) as acquired for the pan-cancer cohort are indicated. Note that the correlations are highest between CD8^+^, PD-1^+^ and CD8^+^PD-1^+^ cells, lower between these cell types and FOXP3^+^ cells, and lowest between these cell types and PD-L1+ cells (tumor only= *tps*, immune cells and tumor cells= *cps*).
